# Evaluating methodology to infer the effect direction in genetic association studies: applications to Body Mass Index, depression, and asthma

**DOI:** 10.64898/2026.08.05.26359780

**Authors:** Tom Chen, Kirsten Voorhies, Adam Reeson, Sujin Seo, Sanghun Lee, Georg Hahn, Julian Hecker, Dmitry Prokopenko, Karin F. Hoth, Rachel S. Kelly, Jessica Lasky Su, Scott T. Weiss, Christoph Lange, Sharon M. Lutz

**Author notes:** Correspondence: Sharon M. Lutz, Department of Population Medicine, Harvard Medical School and Harvard Pilgrim Health Care Institute, 401 Park Dr, Suite 401 East, Boston, MA 02215, USA. These authors contributed equally to this work (Tom Chen, Kirsten Voorhies).

## Abstract

Mendelian Randomization (MR) is a popular tool for inferring causal relationships between traits using genetic variants as instrumental variables. These methods have been extended to also determine the direction of causality (e.g., X → *Y* or *Y* → X). However, causal direction cannot be inferred from a statistical test or estimation procedure (i.e. from data alone) without further assumptions and the methods’ operating characteristics and relative performances are not well understood. We conducted a comprehensive simulation study to illustrate this issue by evaluating type I error and power of 17 summary-based MR methods for inferring the effect direction. These methods fall within three methodological families: MR Steiger, Causal Direction (CD), and bidirectional MR approaches, with scenarios ranging across combinations of horizontal pleiotropy, unmeasured confounding, measurement error, longitudinal feedback, and varying sample sizes. While most methods achieved sufficient power levels under the alternative hypothesis in most scenarios, we found that every method was susceptible to inferring the wrong causal direction or under powered, and no method consistently maintained both correct type 1 error control and high power. In our applications, we evaluated the effect direction between the trait pairs body mass index (BMI) and major depressive disorder (MDD) and between BMI and asthma. To help researchers to evaluate the 17 methods to infer the effect direction and consider these challenges in their own data, we have developed **MRdirection**, an R package that runs the simulation studies examining the 17 directional MR methods across different user-defined scenarios. Our study, together with the accompanying R package, provides researchers with a tool for examining directional MR methods given different underlying assumptions.

## 1. Introduction

Mendelian Randomization (MR) is an increasingly popular method that uses genetic variants as instrumental variables (IVs) to estimate the causal effect of an exposure on an outcome in the presence of unmeasured confounding.[^1, 2^] A fundamental challenge in cross-sectional studies is the possibility of reverse causation. For example, observational cross-sectional studies that have applied univariable MR have consistently reported a strong association between body mass index (BMI) and depression [^3, 4^], with univariable MR concluding that BMI has a causal effect on multiple depression outcomes.[^5^] However, bidirectional MR studies, which attempt to infer the defect direction between two traits, yield mixed results. One study implemented four methods (Inverse-Variance Weighted [IVW], weighted median, Mendelian Randomization Pleiotropy RESidual Sum and Outlier [MR-PRESSO], and Generalized Summary Data-Based Mendelian Randomization [GSMR]) and reported a bidirectional relationship between BMI and depression, while MR-Egger reported no effect in either direction.[^6^] Another study used three methods (IVW, weighted median, MR-Egger) reporting an effect of BMI on depressive symptoms but no reverse effect.[^7^] For BMI and asthma, large-scale Generalized Summary-data-based Mendelian Randomization (GSMR) and Latent Causal Variable (LCV) analyses support an effect of BMI on asthma with little evidence in the reverse direction, whereas a separate two-sample IVW study reported a modest association from allergic asthma to obesity.[^8, 9^] These inconsistent findings point to the need for a clearer understanding of how methodological choices can influence conclusions about causal direction.

Several methods have been proposed that try to infer the direction of causal effects between cross-sectionally measured phenotypes in MR studies. Such methods primarily fall into three classes: MR Steiger, Causal Direction (CD) methods, and Bidirectional MR. Although each of these method classes has been examined before, existing evaluations are limited in scope and across the differing methods. Papers introducing a new directional estimator typically benchmark it against only a few competitors, most often MR Steiger and bidirectional MR, under simulation conditions chosen by the method’s developers (1-3). Broader tutorial reviews of MR catalog these methods and their assumptions but do not evaluate their operating characteristics head-to-head under a common data-generating model (4, 5). Applied studies, meanwhile, routinely use Steiger filtering as directional checks without characterizing how reliable those checks are under certain assumptions (6, 7). Consequently, investigators lack guidance on how the three families compare, how they degrade under realistic violations such as horizontal pleiotropy, unmeasured confounding, measurement error, and longitudinal feedback, and whether their errors surface as spurious reverse-direction calls or as loss of power. To our knowledge, no prior study has evaluated all three families of summary-based directional methods within a single unified simulation framework while jointly reporting type I error, power, and the rate of incorrect direction calls. We address this gap and translate the findings into practical, assumption-aware guidance for applied MR.

The remainder of this paper is organized as follows. In Section 2, we describe these three major classes of methods in detail, outlining the 17 specific approaches evaluated in this study. Section 3 details our simulation study design, including the data-generating mechanisms for Bernoulli and normally distributed phenotypes, the specific scenarios tested (pleiotropy, unmeasured confounding, measurement error, and feedback loops), and the two sample sizes considered (N=1,000 and N=100,000). In Section 4, we present the results of our comprehensive simulations, evaluating each method’s performance on type I error control, power, and its ability to correctly identify causal direction. Section 5 presents four empirical analyses of BMI with major depressive disorder (MDD) and with asthma, using different discovery and target genome wide association study (GWAS) data sources. Finally, Section 6 concludes with a discussion of our findings and their implications for practice. To address this, we provide a comprehensive benchmark of 17 summary-based directional MR methods spanning all three families, characterizing when each method correctly recovers the causal direction and when it fails across a common set of systematically varied assumption violations, and we release the accompanying MRdirection R package so that investigators can reproduce and extend these evaluations in their own settings.

## 2. Material and Methods

We describe three classes of methods for inferring the causal direction using SNPs as IVs: MR Steiger, Causal Direction (CD), and Bidirectional MR. The core statistical assumptions for each method are summarized in Table 1.

**Table 1.** Assumptions of the various methods.

| Category | Method | Estimator | Assumes no horizontal pleiotropy? | Assumes balanced pleiotropy? | Assumes InSIDE? | IV validity assumption (All, Majority, Plurality, None) | Assumes NOME? | Assumes no unmeasured confounding? | Assumes no SNP measurement error? | Assumes phenotypes' measurement errors are uncorrelated? | Assumes no selection bias? |
| --- | --- | --- | --- | --- | --- | --- | --- | --- | --- | --- | --- |
| MR Steiger | MR Steiger IVW (fixed) | IVW | Yes | Yes | Yes | All | Yes | Yes | Yes | Yes | Yes |
|  | MR Steiger Weighted Median | Median | No | No | No | Majority | Yes | Yes | Yes | Yes | Yes |
|  | MR Steiger Egger | Egger | No | No | Yes | None | Yes | Yes | Yes | Yes | Yes |
| CD | CD-Ratio (fixed) | IVW | Yes | Yes | Yes | All | Yes | Yes | Yes | Yes | Yes |
|  | CD-Egger | Egger | No | No | Yes | None | Yes | Yes | Yes | Yes | Yes |
|  | CD-GLS | Egger/GLS | No | No | Yes | None | Yes | Yes | Yes | Yes | Yes |
|  | CD-cML | cML | No | No | No | Plurality | No | Yes | Yes | Yes | Yes |
|  | MR-cML | cML | No | No | No | Plurality | No | Yes | Yes | Yes | Yes |
| Bidirectional | Bidirectional MR IVW | IVW | Yes | Yes | Yes | All | Yes | Yes | No | No | No |
|  | Bidirectional MR IVW multiplicative RE | IVW | No | Yes | Yes | None | Yes | Yes | No | No | No |
|  | Bidirectional MR IVW fixed effects | IVW | Yes | Yes | Yes | All | Yes | Yes | No | No | No |
|  | Bidirectional MR simple median | Median | No | No | No | Majority | Yes | Yes | No | No | No |
|  | Bidirectional MR weighted median | Median | No | No | No | Majority | Yes | Yes | No | No | No |
|  | Bidirectional MR penalized weighted median | Median | No | No | No | Majority | Yes | Yes | No | No | No |
|  | Bidirectional MR Egger | Egger | No | No | Yes | None | Yes | Yes | No | No | No |
|  | Bidirectional MR Egger bootstrap | Egger | No | No | Yes | None | Yes | Yes | No | No | No |
|  | Bidirectional MR unweighted regression | OLS | Yes | No | Yes | All | Yes | Yes | No | No | No |

### 2.1 MR Steiger

The MR Steiger framework evaluates both the presence and direction of a causal relationship by combining two complementary statistical tests.[^10, 11^] First, a standard MR analysis is conducted to test the null hypothesis of no causal effect between phenotype 1 (X) and phenotype 2 (*Y*), yielding a p-value *p*_MR_. This can be implemented using various estimators that offer different robustness properties: the inverse-variance weighted (IVW) method assumes balanced pleiotropy; the weighted median estimator provides consistent estimates if at least 50% of the weight comes from valid instruments; and MR-Egger regression allows for horizontal pleiotropy through its intercept term.

Second, Steiger’s Z-test for dependent correlations is applied to determine whether SNP *g* explains more variance in the exposure or in the outcome. The test compares 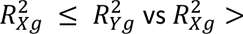 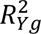, where 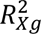 and 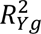represent the correlation between Xand g and the correlation between *Y*and g, respectively. The sign of the test statistic *Z* indicates which trait is more strongly influenced by the genetic instruments, with *Z* > 0 supporting X → *Y* and *Z* < 0 supporting *Y* → X. The corresponding p-value *p*_Steiger_ reflects the significance of this difference. At a chosen significance level *α*, the results are combined to distinguish between three scenarios:

Case 1: If *p*_MR_ < *α*, *p*_Steiger_ < *α*, *Z* > 0, then conclude X causes *Y*.

Case 2: If *p*_MR_ < *α*, *p*_Steiger_ < *α*, *Z* < 0, then conclude *Y* causes X.

Case 3: If *p*_MR_ > *α* or *p*_Steiger_ > *α*, then neither model is accepted.

#### Assumptions

Because MR Steiger relies both on standard MR estimation and on correlation contrasts, its validity rests on a broader set of assumptions. The foundation is the set of *instrumental variable (IV) validity assumptions*, namely instrument relevance (SNPs must be associated with the exposure), independence (instruments must be independent of confounders of the exposure–outcome relationship), and the exclusion restriction, often described in MR as no horizontal pleiotropy (instruments affect the outcome only through the exposure). Different estimators then relax or refine these conditions: the weighted median requires that at least half of the total weight derives from valid instruments; MR-Egger relaxes the exclusion restriction but requires the InSIDE assumption (instrument strength independent of pleiotropic effects); and both IVW and MR-Egger typically assume the NOME condition (no substantial measurement error in SNP–exposure associations). Beyond these MR-specific refinements, the Steiger Z-test introduces additional requirements. Valid inference requires no unmeasured confounding between X and *Y*; no measurement error in SNPs; no horizontal pleiotropy, uncorrelated measurement error across X and *Y* , and absence of selection bias. These extra assumptions are needed because the Steiger test directly compares the squared correlation between the SNP *g* and the phenotype 1 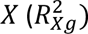 and the squared correlation between the SNP *g* and the phenotype 2 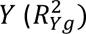. If measurement error is correlated across phenotypes, or if confounding or selection bias induces spurious covariance between SNPs and phenotypes, the relative *R*^2^ can be distorted. Such distortions inflate or deflate the correlation between the SNP and the phenotype, leading the test to infer the wrong causal direction even if the MR estimators themselves remain unbiased.

### 2.2 Causal Direction Methods

Following Xue and Pan’s correlation-based approach to bidirectional MR, we refer to their family of methods as Causal Direction (CD) methods. Introduced and extended across two papers, these methods infer direction by contrasting SNP-phenotype correlations rather than MR effect sizes, encompassing CD-Ratio, CD-Egger, CD-GLS[^12^] and the constrained-maximum-likelihood extensions CD-cML and MR-cML.[^13^] Like MR Steiger, the CD framework models the relationship between SNP-phenotype correlations across variants. But rather than combining an MR estimate with a separate Z-test, CD methods analyze the correlation pairs *R_X_*_g_, *R*_Yg_ across SNPs to infer whether genetic instruments explain more variation in phenotype X or phenotype *Y*. In the simplest case, the CD-Ratio method computes the ratio *R*_Yg_/*R_X_*_g_ (or its reciprocal) for each SNP, making it a direct analogue of the inverse-variance weighted (IVW) estimator in correlation space. These ratios are then aggregated across SNPs to obtain directionality estimates *̂K*_*XY*_ and *̂K*_*X*_. Extensions parallel familiar MR estimators: CD-Egger introduces an intercept term when regressing *R*_fg_ on *R_X_*_g_, allowing for horizontal pleiotropy under the InSIDE assumption, analogous to MR-Egger; CD-GLS improves on this by modeling the joint uncertainty in both *R_X_*_g_ and *R*_Yg_, yielding more efficient inference when multiple variants are analyzed; and CD-cML jointly estimates *̂K_XY_* and *̂K_YX_* within a likelihood framework that accommodates invalid instruments, relying only on the weaker plurality-valid condition. CD-cML’s counterpart, MR-cML, uses the same machinery within the standard MR framework.

As with MR Steiger, the CD methods jointly assess both the existence and direction of a causal relationship, distinguishing among the same three cases as mentioned above. At significance level *α*, we construct confidence intervals 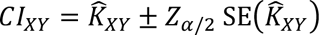 and 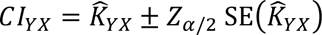 and follow the decision rules:

Case 1: If *CI_XY_* lies entirely outside the no-direction range [−1,1], while *CI_YX_* lies entirely inside [−1,1] and excludes 0, then conclude X causes *Y*.

Case 2: If *CI_YX_* lies entirely outside the no-direction range [−1,1], while *CI_XY_* lies entirely inside [−1,1] and excludes 0, then conclude *Y* causes X.

Case 3: In all other cases, neither model is accepted.

For MR-cML, the decision rules are expressed directly in terms of p-values for each direction. Let *̂θ*_*YX*_ and *̂θ*_*XY*_ denote the estimated causal effects for X → *Y* and *Y* → X, respectively, with corresponding *p*-values *p*_f_*_X_* and *p_X_*_f_. At significance level *α*, the decision rules are:

Case 1: If *p_YX_* < *α* and *p_X_*_Y_ ≥ *α*, then conclude X causes *Y*.

Case 2: If *p_YX_* ≥ *α* and *p_X_*_Y_ < *α*, then conclude *Y* causes X.

Case 3: In all other cases, then neither model is accepted.

#### Assumptions

The validity of CD methods rests on both standard IV assumptions and correlation-based requirements. As above, core IV assumptions include instrument relevance, independence from confounders, and no horizontal pleiotropy. Estimator-specific refinements apply: CD-Ratio assumes all instruments are valid; CD-Egger relaxes the exclusion restriction but requires the InSIDE condition; CD-GLS accounts for variation in both *R_X_*_g_ and *R*_Yg_ but also relies on InSIDE; and CD-cML and MR-cML require the plurality-valid condition. As with IVW and Egger, NOME is typically assumed. Beyond these MR-related assumptions, CD methods also require no unmeasured confounding between X and *Y*, no SNP measurement error, uncorrelated measurement error across phenotypes, and absence of selection bias. Because CD inference compares the relative magnitudes of *R_X_*_g_/*R*_Yg_ and *R*_Yg_/*R_X_*_g_, any distortion in these correlations due to error, confounding, or selection can lead to incorrect inference.

### 2.3 Bidirectional MR

This approach requires two non-overlapping sets of valid instruments: one strongly associated with X, and another strongly associated with *Y*. Standard MR estimation is then performed twice, once treating X as the exposure and *Y* as the outcome, and once with the roles reversed.[^14^] At significance level *α*, let *p_YX_* denote the p-value for the test of X → *Y* and *p_X_*_X_ the p-value for the test of *Y* → X. The decision rules are:

Case 1: If *p_YX_* < *α* and *p_X_*_Y_ ≥ *α*, then conclude X causes *Y*.

Case 2: If *p_YX_* ≥ *α* and *p_X_*_Y_ < *α*, then conclude *Y* causes *X*.

Case 3: In all other cases, the neither model is accepted.

In practice, a range of estimators can be used in the bidirectional framework. We consider bidirectional MR with IVW (including fixed-effects and multiplicative random-effects variants),[^15^] simple median,[^16^] weighted median,[^17^] penalized weighted median, MR-Egger (with and without bootstrapped errors),[^18, 19^] and unweighted regression.[^20^] Each carries its own robustness properties: IVW methods assume no or balanced pleiotropy; median-based methods remain valid if a majority of instruments are valid; and MR-Egger relaxes the exclusion restriction under the InSIDE assumption.[^21^] Unweighted regression, though rarely applied, provides a simple baseline for comparison.

#### Assumptions

Bidirectional MR requires two independent sets of SNPs, one for X and one for *Y*. Each set must satisfy the standard IV validity conditions, but additional considerations arise because inference is performed in both directions. Both sets of SNPs must be sufficiently strong to avoid weak-instrument bias, as instability in either direction can distort conclusions. The two SNP sets should be non-overlapping, since shared SNPs would undermine the ability to distinguish causal direction. Moreover, the two phenotype samples should ideally be independent to minimize bias from sample overlap and inflated SNP–trait associations due to selection in the same dataset. Unlike MR Steiger or CD methods, bidirectional MR does not rely on comparing *R*^2^ values, but its reliability depends on the strength, independence, and separation of the two SNP sets.

## 3. Simulation Setup

### 3.1 Data Generation Framework

We conducted simulation studies to evaluate the performance of MR direction methods under a variety of conditions that relax standard IV assumptions. In what follows, we let Φ(⋅) denote the identity link function for a normally distributed trait and the logistic link function for a binary trait. For each simulation replicate, we generated phenotypes X and *Y* for a sample of N individuals based off the following models:

1. Two independent SNP sets *G_X_* and *G*_f_ are generated from a binomial distribution with size 2 and probability *p* = 0.5 to represent additive coding.
2. The latent (“true”) exposure X^∗^ is generated according to Φ(E[X^∗^]) = *γ_G_G_X_* + *γ_G_*_Y_*G*_Y_ + *γ_U_U*.
3. The measured exposure X equals X^∗^ when there is no measurement error; otherwise, we generate according to Φ(E[X]) = *δ_X_*X^∗^.
4. The outcome *Y* is generated from Φ(E[Y]) = *β_X_*X + *β_G_*_X_ *G_X_* + *β_G_*_Y_ *G*_f_ + *β_U_U*.
5. The unmeasured confounder *U* is generated from E[U] = *η_G_*_X_*G_X_* + *η_G_*_Y_ *G*_f_.
6. If the longitudinal feedback scenario is present, we generate phenotypes to mimic the relation X_1_ → *Y*_1_ → X_2_ → *Y*_2_ as follows: Φ(E[X_2_]) = *κ*_Y_*Y*_1_ and Φ(E[Y_2_]) = *ι_X_*X_2_.

This model is deliberately over-parameterized: not all terms are active in every simulation. Each of the below scenarios and settings corresponds to different choices of parameters in the generative model.

### 3.2 Core Scenarios

We considered 8 baseline scenarios:

- Two sample sizes/genetic effect sizes:

- Scenarios 1 – 4: N = 1,000 with 10 SNPs with genetic effects (*γ_G_*_X_ = *β_G_*_Y_ = 1.0). The four phenotype combinations are both phenotypes are continuously distributed, both phenotypes are binary, phenotype 1 is continuous while phenotype 2 is binary, and phenotype 1 is binary while phenotype 2 is continuous.
- Scenarios 5 – 8: N = 100,000 with 50 SNPs with *γ_G_*_X_ = *β_G_*_Y_ = 0.2. The four phenotype combinations are the same as scenarios 1-4 with both continuous, both binary, or a mix.

### 3.3 Sub-scenarios to Test Violations of Assumptions

For each of the 8 baseline scenarios, we generated the data under the following 6 additional sub scenarios (A-F) (Figure 1). For scenarios B-E, pleiotropy, unmeasured confounding, and/or measurement error are generated. For scenario B and D, 3 types of pleiotropy and unmeasured confounding are considered, respectively. For scenario C, 2 types of measurement error are considered. This results in 8 baseline scenarios and 11 sub scenarios, which results in a total of 88 simulations studies.

A. **Null (Baseline)**: *γ_G_*_Y_ = *β_G_*_X_ = *γ_U_* = *β_U_* = *η_G_*_X_ = *η_G_*_Y_ = 0 and X = X^∗^, meaning that there is no measurement error, no un measured confounding, and no horizontal pleiotropy. Unless otherwise specified, these parameters are assumed to be zero in the subsequent settings.
B. **Pleiotropy**. Nonzero direct SNP effects across traits, with (*γ_G_*_Y_ , *β_G_*_X_ ) = (0, 0.2), (0.2, 0), (0.2, 0.2) for scenarios 5-8 (sample size of N=100,000) and (*γ_G_*_Y_ , *β_G_*_X_ ) = (0, 1), (1, 0), (1, 1) for scenarios 1-4 (sample size of N=1,000).
C. **Measurement Error.** Observed phenotype X measured with reduced reliability *δ_X_* = 0.5 and then *δ_X_* = 1.0 across all 8 scenarios.
D. **Unmeasured Confounding.** Unmeasured confounder *U* influences both X^∗^ and *Y* under configu-rations (*γ_U_*, *β_U_*, *η_G_*_X_ , *η_G_*_Y_ ) = (1,1,0,0), (1, −1,0,0), and (1, 1, 0, 0.2) (for large-sample size scenarios 5-8) / (1, 1, 0, 1) (for small sample size scenarios 1-4).
E. **Combined Violations**. Simultaneously incorporate pleiotropy, measurement error, and unmeasured confounding with *β_G_*_X_ = 0.2 (scenarios 5-8) or *β_G_*_X_ = 1 (scenarios 1-4); *δ_X_* = 1; and *γ_U_* = 1, *β_U_* = −1, respectively.
F. **Feedback Loop.** A longitudinal process where X_1_ → *Y*_1_ → X_2_ → *Y*_2_. Final traits were generated as Φ(X_2_) = *κ*_Y_*Y*_1_ and Φ(*Y*_2_) = *ι_X_*X_2_ with *κ*_Y_ = 0.2 and *ι_X_* = 0.2.

**Figure 1.**
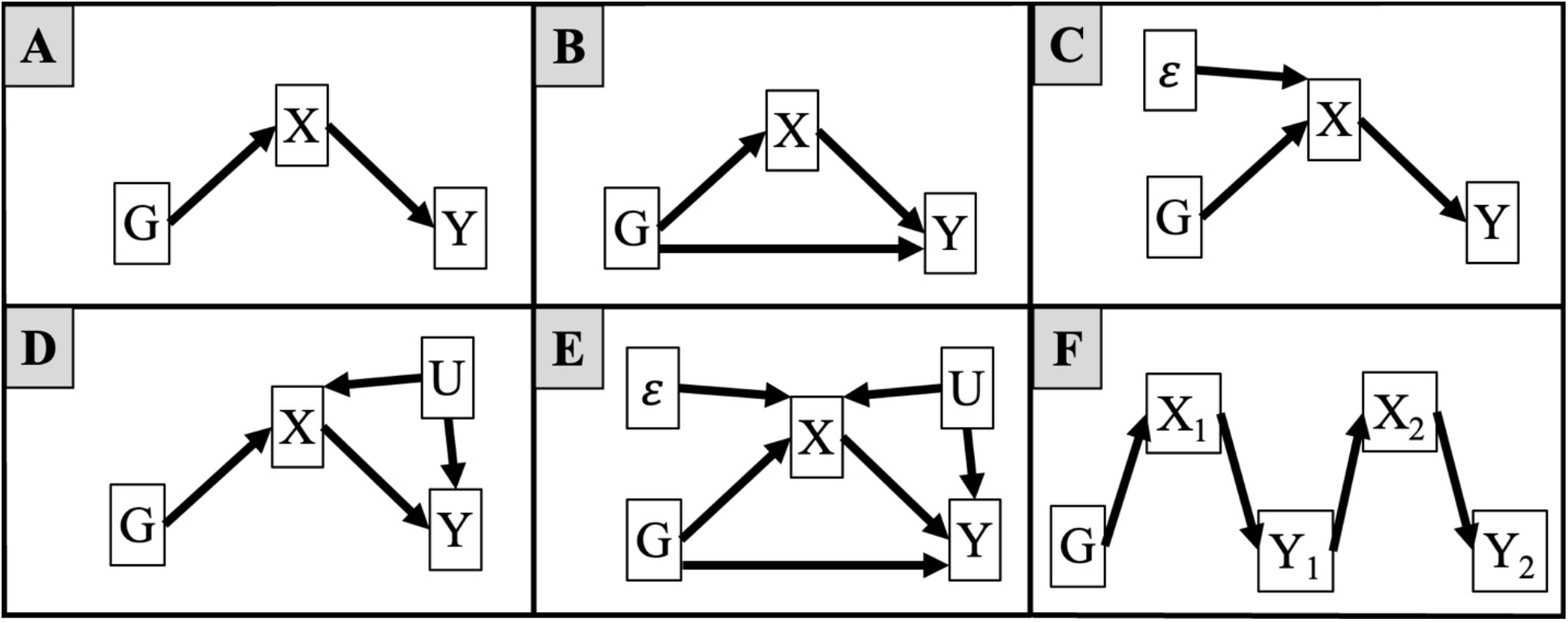
Directed acyclic graphs of sub scenarios A-F. A is the scenario that has no pleiotropy, no measurement error, and no unmeasured confounding. B is the scenario with pleiotropy. C is the scenario with measurement error. D is the scenario with unmeasured confounding, E is the scenario with pleiotropy, measurement error, and unmeasured confounding. F is the scenario with a longitudinal feedback loop.

## 4. Simulation Results

We evaluated 17 MR methods to infer the effect direction across 8 core scenarios and 6 sub scenarios (A–F). Each scenario was replicated 1,000 times and analyses were performed at a significance level of *α* = 0.05. Performance was summarized in terms of type I error control (with >10% considered substantially inflated), the frequency of incorrectly inferring reverse causation (Case 2: *Y* → X, with >5% considered spurious), and the frequency of correctly detecting the true direction (Case 1: X → *Y*, with <75% indicating low power). Full results for Scenario 5 (large sample, both traits continuous) are shown in Table 2 and Figure 2, with complete results for all other scenarios and sub scenarios provided in Supplementary Tables S1–S7 and Figures S1–S7.

**Figure 2.**
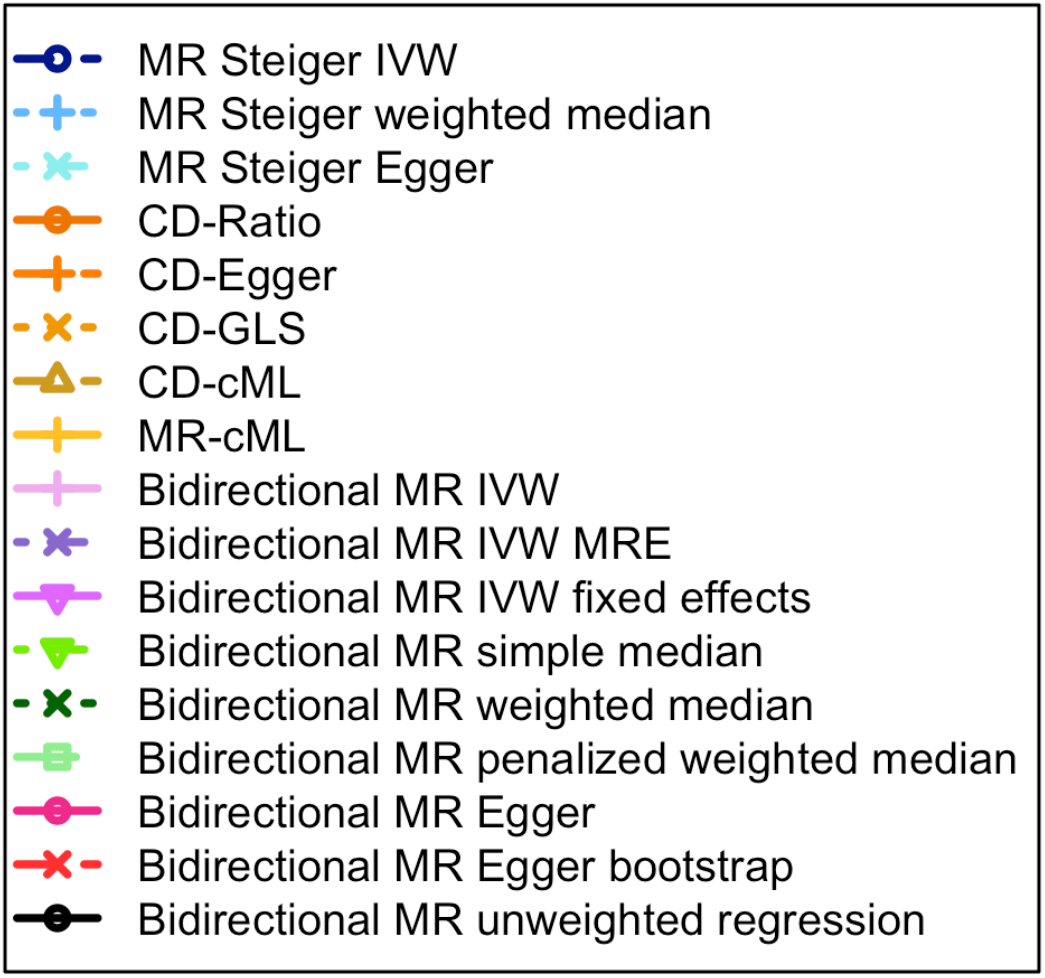
Simulation results for Scenario 5 (N = 100,000, 50 SNPs, minor allele frequency = 0.5, *γ_GX_* = *β_G_*_Y_ = 0.2). Both phenotypes are normally distributed. Columns correspond to sub-scenarios (A–F); rows show the proportion of simulations returning case 1 (X → *Y*, top) or case 2 (*Y* → X, bottom) across values of *β_X_*. The x-axis shows the true effect size *β_X_* and the y-axis shows the proportion of simulations where each method returned the given case. Line colors correspond to MR methods (see legend). Note MR IVW MRE stands for Inverse Variance Weighted (IVW) method with a Random Effects (MRE) model.

**Table 2.**
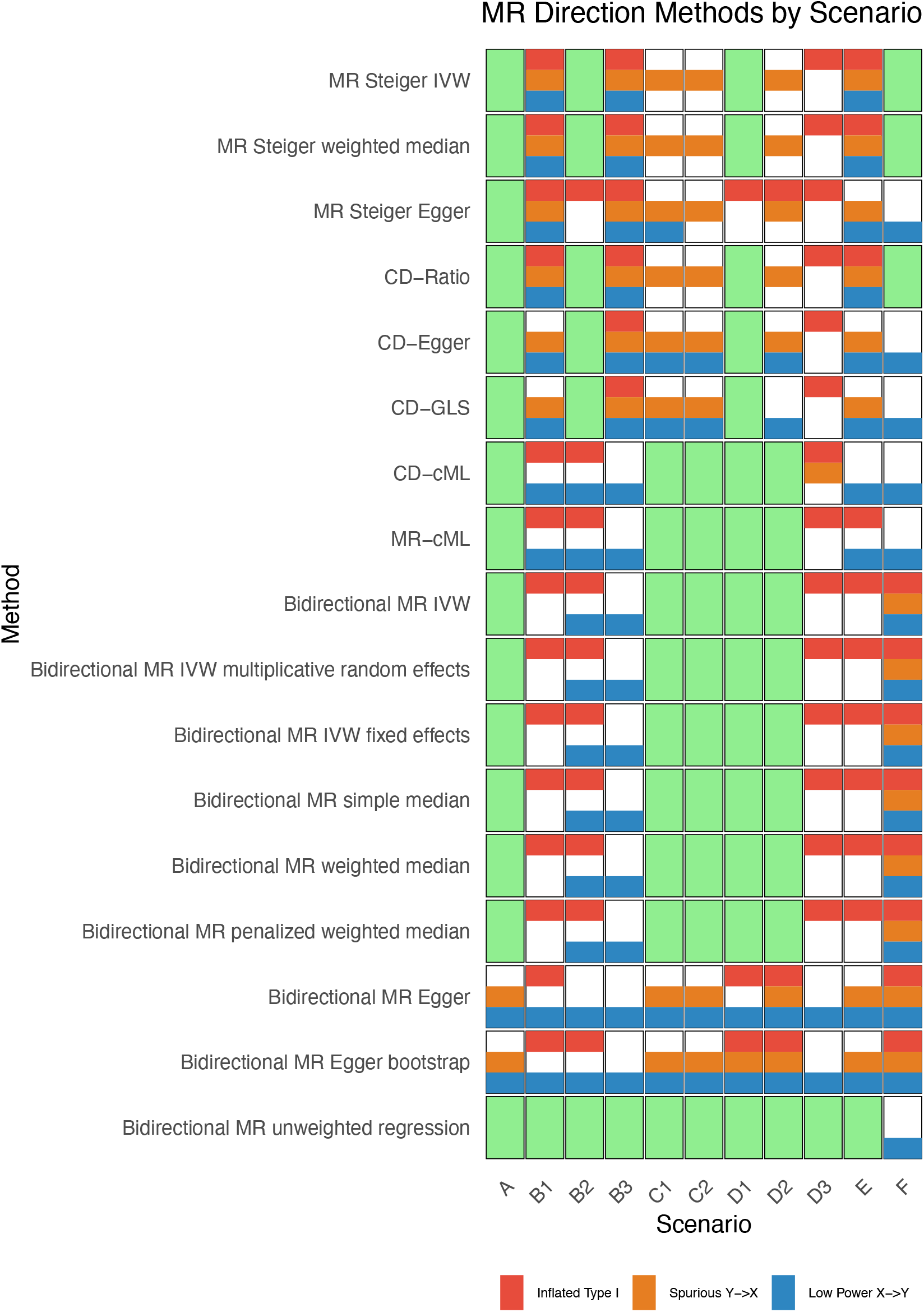
Simulation results with *N* = 100,000 and 50 SNPs (Y*_GX_* = 0.2, *β_G_*_Y,_ = 0.2a). Heat maps summarize performance across the range of *β_X_* values shown in Figure 2. Shading indicates where methods showed inflated Type I error (>10%, red), spurious *Y* → *X* inference (>5%, orange), or low power to detect *X* → *Y* (<75%, blue) in at least one setting. Cells without shading indicate consistently high performance across all checks.

### 4.1 Type I Error Performance

Methods that rely on the assumption of valid instruments (no horizontal pleiotropy) were highly sensitive when this condition was violated. MR Steiger IVW, MR Steiger weighted median, and CD-Ratio showed nearly complete type I error inflation when *G_X_* were pleiotropic (B1) or when both instrument sets were pleiotropic (B3). The same pattern was observed for the bidirectional MR IVW and median estimators, which also require valid instruments in each direction. These effects were more pronounced in the larger-sample scenarios (5–8), where small violations were more detectable and translated into systematic misclassification.

Unmeasured confounding, especially when genetic instruments affected the confounder (D3), produced elevated Type I error rates for most methods, including Steiger procedures, CD-cML, MR-cML, and bidirectional MR estimators. Milder confounding (D1–D2) also led to inflation for correlation-based methods such as Steiger-Egger, CD-Egger, and CD-GLS. The combined violation scenario (E) resulted in high error rates across nearly all methods. Measurement error alone (C1–C2) had limited impact on IVW or median approaches but reduced reliability for Steiger and CD methods, which depend on contrasts of explained variance.

Bidirectional MR estimators did not rely on *R*^2^ contrasts and were less affected by measurement error. Their main limitations were consistent inflation under pleiotropy, confounding where *G_X_* influenced the confounder (D3), and the feedback loop (F), which created genuine bidirectionality. The bidirectional MR Egger estimators showed the broadest range of inflated error, with failures across most pleiotropy and confounding scenarios as well as under feedback. The unweighted regression method maintained nominal Type I error across all settings, reflecting its reduced sensitivity to individual pleiotropic or confounded variants when their effects were dispersed, though this came at the cost of efficiency relative to other estimators.

### 4.2 Incorrect Detection of *Y* → X

Methods relying on comparisons of genetic correlation strength (*R*²), including the MR Steiger framework and several CD methods, were particularly susceptible to spurious *Y* → X calls when the relative variance explained by instruments was distorted. This occurred systematically under measurement error scenarios (C1, C2), where correlated error across phenotypes artificially inflates the apparent genetic correlation with the outcome. Similarly, unmeasured confounding (particularly D2, D3) and combined violations (E) induced *Y* → X causation by creating pathways that distorted correlation comparisons.

The constrained maximum likelihood methods (CD-cML, MR-cML) demonstrated greater robustness in larger samples (N=100,000), with MR-cML showing no spurious *Y* → X calls in these scenarios. However, both methods showed increased susceptibility in smaller samples (N=1,000), where estimation uncertainty is greater.

In contrast, the bidirectional MR methods (IVW, median variants) demonstrated substantially better control of spurious *Y* → X inference, consistent with their independence from correlation-based assumptions. Spurious calls emerged in the presence of a true feedback loop (F), where the causal model itself violates the assumption of unidirectionality. The bidirectional MR Egger methods constituted an exception to this pattern, exhibiting widespread spurious *Y* → X calls across most violation scenarios, suggesting limitations in their directional robustness within the bidirectional framework. Bidirectional MR with the unweighted regression method was the most reliable, with spurious *Y* → X calls occurring only in small-sample, strong-effect scenarios when pleiotropy or feedback was present.

### 4.3 Correct Detection of X → *Y*

The bidirectional MR methods (IVW and median variants) demonstrated the most consistent performance for correct detection. These methods maintained high power under the null scenario (A), measurement error (C1, C2), and unmeasured confounding (D1, D2) across most sample sizes and trait types. This robustness is consistent with their design, which does not rely on comparing genetic correlation strengths and is therefore robust to the distortions that affect correlation-based methods under these conditions. Their performance was more variable under pleiotropic scenarios (B1, B2), which directly challenge their requirement for valid, exposure-specific instruments.

The correlation-based methods, including the MR Steiger framework and several CD estimators, showed more constrained performance. They achieved correct detection primarily in scenarios with larger genetic effects (Scenarios 5-8) and under limited violations, such as specific pleiotropic conditions (B2) or simple unmeasured confounding (D1, D2). Their performance was generally compromised in smaller samples (N=1,000) and under violations that distort genetic correlations, such as measurement error (C1, C2) or complex confounding (D3, E). This pattern underscores their dependence on the accurate estimation of relative variance explained (*R*²), which is sensitive to these violations.

The constrained maximum likelihood methods (CD-cML, MR-cML) exhibited a strong sample size dependency. They rarely achieved the performance benchmark in smaller samples but showed improved correct detection in larger samples (N=100,000) under the null scenario (A) and some violation scenarios (D1, D2, C2). This suggests that their model-based approach to handling invalid instruments requires substantial statistical power for reliable performance.

Neither the bidirectional MR Egger methods nor several CD-Egger variants achieved the benchmark for correct detection under any scenario, indicating fundamental limitations in their power or stability for directional inference in this context.

## 5. Application

We applied 17 MR methods to infer the effect direction between 2 phenotypes: BMI⫘MDD (Analyses 1–2) and BMI⫘asthma (Analyses 3–4), using summary statistics from the MRC IEU OpenGWAS infrastructure. Analyses 1–2 differ by the discovery GWAS used to select MDD instruments; Analyses 3–4 differ by the target GWAS used to apply those instruments for asthma. Each analysis was run under two matched SNP thresholds (either p < 5×10⁻⁸ or p < 5×10⁻⁷) applied to both X ⫘ *Y* (i.e., no mixing thresholds within an analysis). For all four analyses, BMI instruments were taken from the GIANT discovery GWAS (n=339,224)[^21^] and applied to BMI from a UK Biobank target GWAS (n = 461,460),[^22^] yielding 78 SNPs at p < 5×10⁻⁸ and 115 SNPs at p < 5×10⁻⁷.

### 5.1 Analysis 1: BMI and MDD with the Psychiatric Genomics Consortium discovery GWAS, UK Biobank target GWAS

Previous studies have supported a causal effect of higher BMI on depression risk[^23, 24^] and mixed evidence for the reverse direction.[^6, 25^] We test these directions in our first and second analyses. At the p < 5×10⁻⁸ threshold, 5 MDD SNPs were selected from a Psychiatric Genomics Consortium (PGC) GWAS (59,851 cases; 113,154 controls) [^26^] and applied to a UK Biobank MDD GWAS (29,475 cases; 63,482 controls).[^27^] Four methods inferred BMI→MDD (Case 1)—MR Steiger IVW, bidirectional MR IVW, bidirectional MR IVW (multiplicative random effects), and bidirectional MR simple median. No method supported MDD→BMI (Case 2); all others were inconclusive (Case 3). At the p < 5×10⁻⁷ threshold, 16 MDD SNPs were selected. Here, additional methods supported BMI→MDD, including CD-cML and MR-cML (all Case 1), while bidirectional MR penalized weighted median suggested MDD→BMI (Case 2); most remaining methods were inconclusive (Case 3). Full results are shown in Table 3.

**Table 3.** Results from 17 directional MR methods for BMI ↔ MDD. X is fixed as BMI (SNPs discovered in GIANT, ieu-a-2, and applied in UK Biobank, ukb-b-19953). For Y (MDD), instruments were discovered either in PGC (ieu-a-1188) or a larger meta-analysis (ieu-b-102) and applied in UK Biobank (ebi-a-GCST009979). Columns show the two discovery sources crossed with instrument thresholds (p < 5×10⁻⁸; p < 5×10⁻⁷). Each cell reports: Case 1 (X→Y), Case 2 (Y→X), or Case 3 (inconclusive).

| Trait pair | BMI MDD |  |  |  |
| --- | --- | --- | --- | --- |
| Threshold | p < 5×10<sup>-8</sup> | p < 5×10<sup>-7</sup> | p < 5×10<sup>-8</sup> | p < 5×10<sup>-7</sup> |
| SNP-discovery source for<br>(OpenGWAS ID) | PGC<br>(ieu-a-1188) |  | Meta-Analysis<br>(ieu-b-102) |  |
| Target source for<br>(OpenGWAS ID) | UK Biobank<br>(ebi-a-GCST009979) |  | UK Biobank<br>(ebi-a-GCST009979) |  |
| Method | Case Returned |  |  |  |
| MR Steiger IVW | 1 | 1 | 1 | 1 |
| MR Steiger weighted median | 3 | 3 | 3 | 3 |
| MR Steiger Egger | 3 | 3 | 3 | 3 |
| CD-Ratio | 3 | 3 | 3 | 3 |
| CD-Egger | 3 | 3 | 3 | 3 |
| CD-GLS | 3 | 3 | 3 | 3 |
| CD-cML | 3 | 1 | 3 | 3 |
| MR-cML | 3 | 1 | 3 | 3 |
| Bidirectional MR IVW | 1 | 1 | 3 | 3 |
| Bidirectional MR IVW multiplicative<br>random effects | 1 | 1 | 3 | 3 |
| Bidirectional MR IVW fixed effects | 3 | 3 | 3 | 3 |
| Bidirectional MR simple median | 1 | 3 | 3 | 2 |
| Bidirectional MR weighted median | 3 | 3 | 2 | 2 |
| Bidirectional MR penalized weighted<br>median | 3 | 2 | 2 | 2 |
| Bidirectional MR Egger | 3 | 3 | 3 | 3 |
| Bidirectional MR Egger bootstrap | 3 | 3 | 2 | 3 |
| Bidirectional MR unweighted regression | 3 | 3 | 3 | 3 |

### 5.2 Analysis 2: BMI and MDD with a meta-analysis discovery GWAS, UK Biobank target GWAS

To increase instrument yield and power, we repeated the analysis using MDD instruments from a larger meta-analysis (47 SNPs at *p* < 5×10⁻⁸; 87 SNPs at *p* < 5×10⁻⁷; 170,756 cases; 329,443 controls),[^28^] applied to the same UK Biobank MDD outcome. At p < 5×10⁻⁸, most methods were inconclusive (Case 3); BMI→MDD (Case 1) was supported by MR Steiger IVW, while MDD→BMI (Case 2) appeared for the bidirectional MR weighted median, penalized weighted median, and Egger bootstrap. At p < 5×10⁻⁷, most methods again returned Case 3; Case 1 was supported by MR Steiger IVW, whereas Case 2 was suggested by the bidirectional MR simple median, weighted median, and penalized weighted median. Overall, we see that direction estimates can shift with choice in discovery GWAS. Full results are shown in Table 3.

### 5.3 Analysis 3: BMI and asthma with a broad-definition asthma, FinnGen discovery GWAS, UK Biobank target GWAS

MR studies typically find that higher BMI increases asthma risk, while evidence for asthma influencing BMI is limited.[^8, 9^] To assess both directions with maximal outcome power, we applied FinnGen-derived asthma instruments (20,629 cases; 135,449 controls)[^29^] to a broad UK Biobank asthma phenotype coded by aggregating self-report, hospital ICD-9/10, and primary-care record review (56,167 cases; 352,255 controls).[^30^] A total of 16 BMI SNPs (at *p* < 5×10⁻⁸) and 24 asthma SNPs (at *p* < 5×10⁻⁷) remained. At both thresholds, results coincided: 11 methods supported BMI→asthma (Case 1) and 6 were inconclusive (Case 3); none supported asthma→BMI (Case 2). Full results are in Table 4.

**Table 4.** Results from 17 directional MR methods for BMI ↔ asthma. X is fixed as BMI (SNPs discovered in GIANT, ieu-a-2, and applied in UK Biobank, ukb-b-19953). For Y (asthma), instruments were discovered in FinnGen (finn-b-J10_ASTHMA) and applied to two UK Biobank targets: a broad asthma phenotype (ebi-a-GCST90014325) and a narrow inpatient ICD-10 J45 phenotype (ukb-d-J45). Columns show the two targets crossed with instrument thresholds (p < 5×10⁻⁸; p < 5×10⁻⁷). Each cell reports: Case 1 (X→Y), Case 2 (Y→X), or Case 3 (inconclusive).

|  | BMI asthma |  |  |  |
| --- | --- | --- | --- | --- |
| Threshold | p < 5×10<sup>-8</sup> | p < 5×10<sup>-7</sup> | p < 5×10<sup>-8</sup> | p < 5×10<sup>-7</sup> |
| SNP-discovery source for<br>(OpenGWAS ID) | FinnGen<br>(finn-b-J10_ASTHMA) |  | FinnGen<br>(finn-b-J10_ASTHMA) |  |
| Target source for<br>(OpenGWAS ID) | UK Biobank<br>(ebi-a-GCST90014325) |  | UK Biobank<br>(ukb-d-J45) |  |
| Method | Case Returned |  |  |  |
| MR Steiger IVW | 1 | 1 | 3 | 3 |
| MR Steiger weighted median | 1 | 1 | 3 | 3 |
| MR Steiger Egger | 3 | 3 | 3 | 3 |
| CD-Ratio | 3 | 3 | 3 | 3 |
| CD-Egger | 1 | 1 | 3 | 3 |
| CD-GLS | 3 | 3 | 3 | 3 |
| CD-cML | 1 | 1 | 3 | 3 |
| MR-cML | 1 | 1 | 3 | 3 |
| Bidirectional MR IVW | 1 | 1 | 3 | 3 |
| Bidirectional MR IVW multiplicative<br>random effects | 1 | 1 | 3 | 3 |
| Bidirectional MR IVW fixed effects | 1 | 1 | 3 | 3 |
| Bidirectional MR simple median | 1 | 1 | 3 | 3 |
| Bidirectional MR weighted median | 1 | 1 | 3 | 3 |
| Bidirectional MR penalized weighted<br>median | 1 | 1 | 3 | 1 |
| Bidirectional MR Egger | 3 | 3 | 2 | 3 |
| Bidirectional MR Egger bootstrap | 3 | 3 | 3 | 3 |
| Bidirectional MR unweighted regression | 3 | 3 | 3 | 3 |

### 5.4 Analysis 4: BMI and asthma with a narrow-definition of asthma, FinnGen discovery GWAS, UK Biobank target GWAS

To assess whether a broad asthma definition diluted signals through phenotype heterogeneity, we repeated the analysis using a stricter UK Biobank inpatient ICD-10 J45 outcome (1,693 cases; 359,501 controls),[^31^] accepting a substantial loss of sample size. Using the same FinnGen instruments, 15 (at p < 5×10⁻⁸) and 23 (at p < 5×10⁻⁷) SNPs remained after harmonization (vs 16/24 in Analysis 3). At p < 5×10⁻⁸, no method supported BMI → asthma (Case 1); one method—bidirectional MR Egger—suggested asthma → BMI (Case 2); the remaining 16 methods were inconclusive (Case 3). At p < 5×10⁻⁷, one method (bidirectional MR penalized weighted median) supported BMI→asthma (Case 1); no methods supported asthma→BMI (Case 2); and 16 methods remained inconclusive (Case 3).

Taking Analysis 3 (broad asthma) and Analysis 4 (narrow asthma) together, the narrow inpatient phenotype improves specificity but reduces precision, pushing most methods to inconclusive calls; isolated findings (Case 2 at 5×10⁻⁸ for MR-Egger; Case 1 at 5×10⁻⁷ for penalized weighted median) should be interpreted cautiously given small case counts and potential estimator sensitivity under weak-instrument conditions. Full results are in Table 4.

## 6. Discussion

Across 17 MR methods to infer the effect direction, no single one consistently delivered reliable directional inference. In simulations spanning pleiotropy, measurement error, unmeasured confounding, and feedback, we found correlation-based approaches (Steiger/CD) failed when relative variance explained (*R*²) was distorted; bidirectional IVW/median estimators were most vulnerable to pleiotropy and to genuine bidirectionality; and cML methods were better able to infer the effect direction with larger sample sizes. These patterns recurred in the applications: for inferring the effect direction between MDD and BMI, increasing the number of SNPs included in the analysis increased the power but also heterogeneity, producing less consistent direction calls. For inferring the effect direction between asthma and BMI, shifting from a broad to a narrow asthma phenotype traded power for specificity and pushed many methods to be inconclusive. Directional conclusions thus depend heavily on method assumptions, SNP discovery sources, and phenotype definitions, and not on method choice alone.

We found that the different method families, likely driven by differences in their core assumptions, had different strengths and weaknesses. Steiger/CD procedures rely on correctly estimating phenotype-specific *R*²; they are most appropriate when measurement error is limited, instruments are comparably measured across phenotypes, and sample overlap and LD differences are minimal, but their performance deteriorates when these conditions are not met. Bidirectional IVW/median methods avoid *R*² comparisons and are therefore less sensitive to measurement error, yet they assume valid, exposure-specific instruments in each direction and are susceptible to horizontal pleiotropy and true bidirectionality. cML approaches can tolerate some invalid instruments but typically require larger sample sizes and careful tuning, and they remain sensitive to complex confounding (e.g., D3). Bidirectional MR-Egger provides a pleiotropy adjustment in principle but showed limited power and stability in our evaluations. Unweighted regression maintained near-nominal Type I error, albeit with reduced efficiency. Based on these results, IVW/median methods are reasonable when pleiotropy is minimal and cML is useful with many instruments and adequate sample size. Steiger/CD should be reserved for settings where *R*² comparability is valid.

This study has several limitations. Despite our best efforts, our applications may involve residual sample overlap between discovery and target GWAS datasets, which can bias summary-based estimators under weak instruments. Phenotype definitions (broad vs narrow asthma) can influence instrument validity and transportability. We did not systematically vary instrument thresholds or LD clumping parameters, nor did we explore winner’s curse in instrument discovery, or ancestry/LD-reference mismatches; each can affect directional calls. Although our simulations spanned multiple violations, they did not include very large samples (>1M), thousands of instruments, or complex multi-ancestry settings since this would be too computationally expensive. We focused exclusively on summary-based methods (excluding individual-level approaches and designs using negative controls or within-family data).

Finally, we caution against vote-counting across methods. The fact that different procedures point in different directions underscores how sensitive conclusions are to method-specific assumptions; our purpose is to characterize that assumption sensitivity, not to aggregate ‘wins.’ To facilitate further investigation, we provide the **MRdirection** R package to reproduce scenarios and extend them to alternative instrument rules, and phenotype definitions, available on GitHub at https://github.com/SharonLutz/MRdirection.

## Data Availability

All data produced are available online at the MRC IEU OpenGWAS repository https://opengwas.io

https://github.com/SharonLutz/MRdirection

## Acknowledgments

This work was supported by NIH grants R01MH129337 and R01HL176792.

## Author contributions

S.M.L, and K.V. created the code for the simulation studies. T.C,, K.V., and S.M.L. conducted the data analyses. S.M.L, K.V., and T.C. drafted the manuscript. T.C., K.V., and A.R. created the plots for the manuscript and supplement. All authors contributed to the editing and drafting of the manuscript.

## Web resources

**MRdirection** is an R package that runs the simulation studies examining the 17 directional MR methods across different user-defined scenarios available on GitHub. https://github.com/SharonLutz/MRdirection

## Data and Code Availability

The code for this study is available via the MRdirection R package on GitHub (https://github.com/SharonLutz/MRdirection). The data analysis used summary statistics from the MRC IEU OpenGWAS repository.

## Declaration of generative AI in scientific writing

Harvard AI Sandbox was used to check for typos and grammar errors in parts on the manuscript.

---

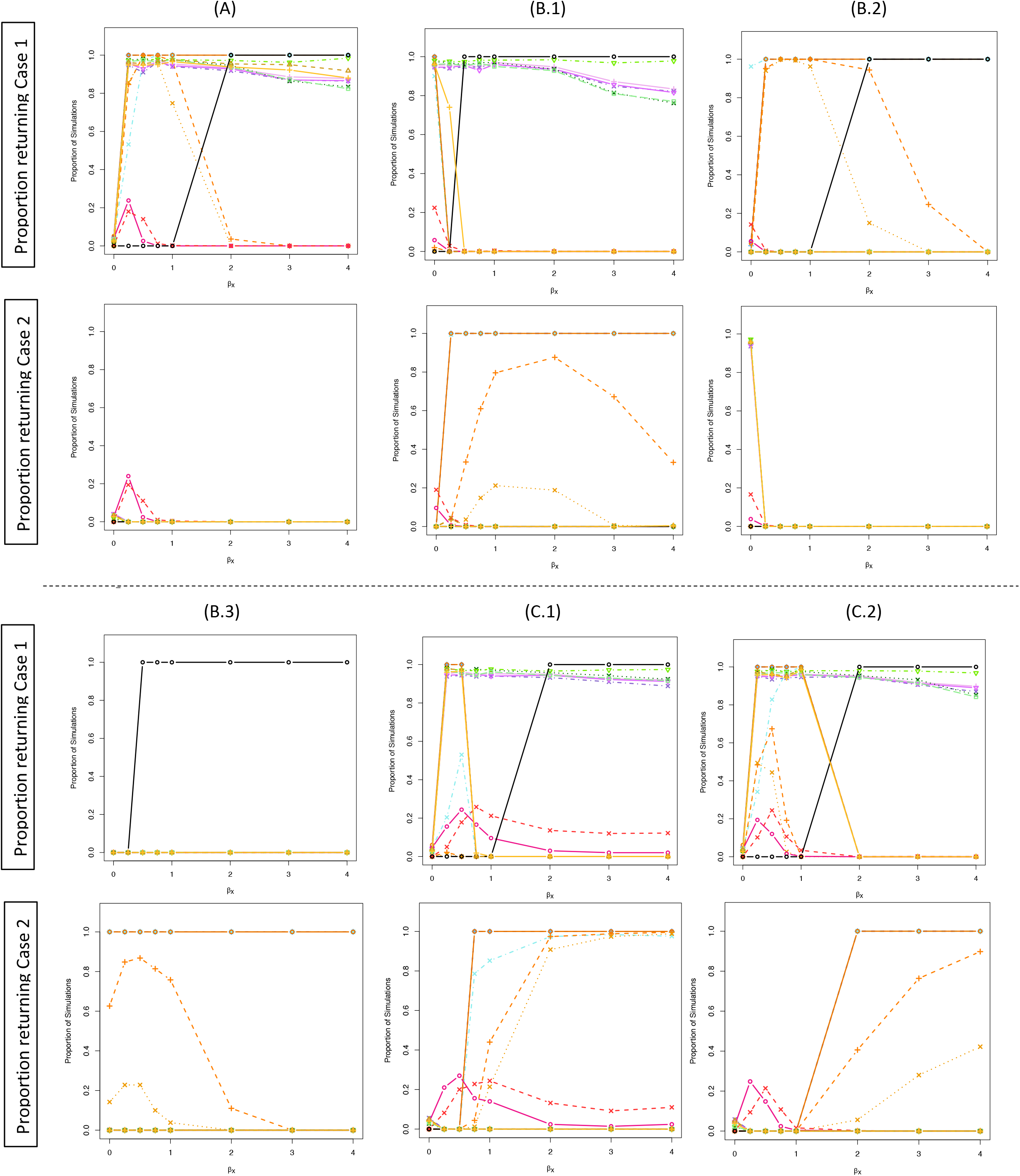
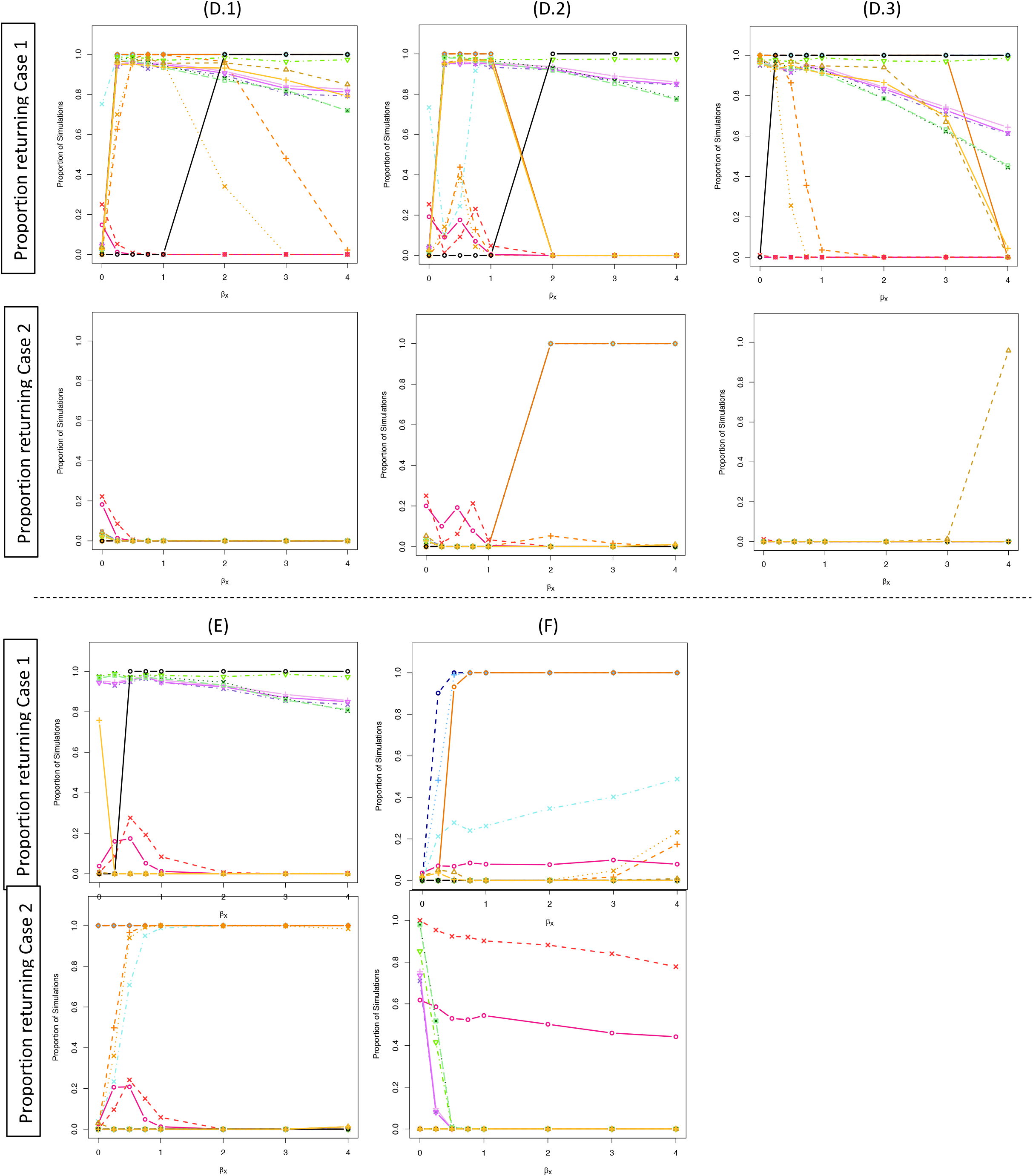

## References

1. Davey Smith G, Ebrahim S. ‘Mendelian randomization’: can genetic epidemiology contribute to understanding environmental determinants of disease? International journal of epidemiology. 2003;32(1):1–22.

2. Zheng J, Baird D, Borges MC, Bowden J, Hemani G, Haycock P, et al. Recent Developments in Mendelian Randomization Studies. Curr Epidemiol Rep. 2017;4(4):330–45.

3. Simon GE, Von Korff M, Saunders K, Miglioretti DL, Crane PK, Van Belle G, et al. Association between obesity and psychiatric disorders in the US adult population. Archives of general psychiatry. 2006;63(7):824–30.

4. Luppino FS, de Wit LM, Bouvy PF, Stijnen T, Cuijpers P, Penninx BW, et al. Overweight, obesity, and depression: a systematic review and meta-analysis of longitudinal studies. Arch Gen Psychiatry. 2010;67(3):220–9.

5. Karageorgiou V, Casanova F, O’Loughlin J, Green H, McKinley TJ, Bowden J, et al. Body mass index and inflammation in depression and treatment-resistant depression: a Mendelian randomisation study. BMC Med. 2023;21(1):355.

6. Zhang H, Zheng R, Yu B, Yu Y, Luo X, Yin S, et al. Dissecting shared genetic architecture between depression and body mass index. BMC medicine. 2024;22(1):455.

7. van den Broek N, Treur JL, Larsen JK, Verhagen M, Verweij KJ, Vink JM. Causal associations between body mass index and mental health: a Mendelian randomisation study. J Epidemiol Community Health. 2018;72(8):708–10.

8. Zhu Z, Guo Y, Shi H, Liu C-L, Panganiban RA, Chung W, et al. Shared genetic and experimental links between obesity-related traits and asthma subtypes in UK Biobank. Journal of Allergy and Clinical Immunology. 2020;145(2):537–49.

9. Li Y, Kan X. Mendelian randomization analysis to analyze the genetic causality between different levels of obesity and different allergic diseases. BMC Pulmonary Medicine. 2023;23(1):352-.

10. Hemani G, Zheng J, Elsworth B, Wade KH, Haberland V, Baird D, et al. The MR-Base platform supports systematic causal inference across the human phenome. elife. 2018;7:e34408.

11. Hemani G, Tilling K, Davey Smith G. Orienting the causal relationship between imprecisely measured traits using GWAS summary data. PLoS genetics. 2017;13(11):e1007081.

12. Xue H, Pan W. Inferring causal direction between two traits in the presence of horizontal pleiotropy with GWAS summary data. PLoS genetics. 2020;16(11):e1009105.

13. Xue H, Pan W. Robust inference of bi-directional causal relationships in presence of correlated pleiotropy with GWAS summary data. PLoS genetics. 2022;18(5):e1010205.

14. Richmond RC, Davey Smith G. Commentary: Orienting causal relationships between two phenotypes using bidirectional Mendelian randomization. International journal of epidemiology. 2019;48(3):907–11.

15. Burgess S, Butterworth A, Thompson SG. Mendelian randomization analysis with multiple genetic variants using summarized data. Genetic epidemiology. 2013;37(7):658–65.

16. Bowden J, Del Greco M F, Minelli C, Davey Smith G, Sheehan N, Thompson J. A framework for the investigation of pleiotropy in two-sample summary data Mendelian randomization. Statistics in medicine. 2017;36(11):1783–802.

17. Bowden J, Davey Smith G, Haycock PC, Burgess S. Consistent estimation in Mendelian randomization with some invalid instruments using a weighted median estimator. Genetic epidemiology. 2016;40(4):304–14.

18. Bowden J, Davey Smith G, Burgess S. Mendelian randomization with invalid instruments: effect estimation and bias detection through Egger regression. International journal of epidemiology. 2015;44(2):512–25.

19. Burgess S, Thompson SG. Interpreting findings from Mendelian randomization using the MR-Egger method. European journal of epidemiology. 2017;32(5):377–89.

20. Unweighted regression - Mendelian randomization dictionary. 2025 Dec 18.

21. Locke AE, Kahali B, Berndt SI, Justice AE, Pers TH, Day FR, et al. Genetic studies of body mass index yield new insights for obesity biology. Nature. 2015;518(7538):197–206.

22. Elsworth B, Mitchell R, Raistrick C, Paternoster L, Hemani G, Gaunt T. Mrc ieu UK Biobank gwas pipeline version 1. University of Bristol. 2017;10.

23. Tyrrell J, Mulugeta A, Wood AR, Zhou A, Beaumont RN, Tuke MA, et al. Using genetics to understand the causal influence of higher BMI on depression. Int J Epidemiol. 2019;48(3):834–48.

24. O’Loughlin J, Casanova F, Fairhurst-Hunter Z, Hughes A, Bowden J, Watkins ER, et al. Mendelian randomisation study of body composition and depression in people of East Asian ancestry highlights potential setting-specific causality. BMC Med. 2023;21(1):37.

25. Mulugeta A, Zhou A, Vimaleswaran KS, Dickson C, Hypponen E. Depression increases the genetic susceptibility to high body mass index: Evidence from UK Biobank. Depress Anxiety. 2019;36(12):1154–62.

26. Wray NR, Ripke S, Mattheisen M, Trzaskowski M, Byrne EM, Abdellaoui A, et al. Genome-wide association analyses identify 44 risk variants and refine the genetic architecture of major depression. Nature genetics. 2018;50(5):668–81.

27. Coleman JR, Peyrot WJ, Purves KL, Davis KA, Rayner C, Choi SW, et al. Genome-wide gene-environment analyses of major depressive disorder and reported lifetime traumatic experiences in UK Biobank. Molecular psychiatry. 2020;25(7):1430–46.

28. Howard DM, Adams MJ, Clarke T-K, Hafferty JD, Gibson J, Shirali M, et al. Genome-wide meta-analysis of depression identifies 102 independent variants and highlights the importance of the prefrontal brain regions. Nature neuroscience. 2019;22(3):343–52.

29. Kurki MI, Karjalainen J, Palta P, Sipila TP, Kristiansson K, Donner KM, et al. FinnGen provides genetic insights from a well-phenotyped isolated population. Nature. 2023;613(7944):508–18.

30. Valette K, Li Z, Bon-Baret V, Chignon A, Berube JC, Eslami A, et al. Prioritization of candidate causal genes for asthma in susceptibility loci derived from UK Biobank. Commun Biol. 2021;4(1):700.

31. Neale L, OpenGWAS MI. Diagnoses – main ICD10: J45 Asthma (UK Biobank). MRC Integrative Epidemiology Unit, University of Bristol; 2018.

