## Supplemental Figures for "Evaluating methodology to infer the effect direction in genetic association studies: applications to Body Mass Index, depression, and asthma"

### Supplementary Materials for "Querying methodology to infer the effect direction in genetic association studies: applications to Body Mass Index, depression, and asthma"

Phenotype 1: Bernoulli distributed; Phenotype 2: normally distributed; sample size $N = 100{,}000$; SNP effect size of 0.2. 15

Phenotype 1: normally distributed; Phenotype 2: Bernoulli distributed; sample size $N = 100{,}000$; SNP effect size of 0.2. 18

Phenotype 1: Bernoulli distributed; Phenotype 2: Bernoulli distributed; sample size $N = 100{,}000$; SNP effect size of 0.2. 21

### Overview of supplemental tables and figures

The supplemental figures display results from the simulation studies under scenarios A–F. Colors indicate method families: MR Steiger (blue), CD methods (orange/yellow/brown), bidirectional MR IVW (purple), bidirectional MR Egger (red/pink), bidirectional MR median (green), and bidirectional MR unweighted regression (black).

In all plots, **Case 1** refers to the method concluding that phenotype 1 causes phenotype 2 ($X \rightarrow Y$), while **Case 2** refers to the method incorrectly concluding that phenotype 2 causes phenotype 1 ($Y \rightarrow X$).

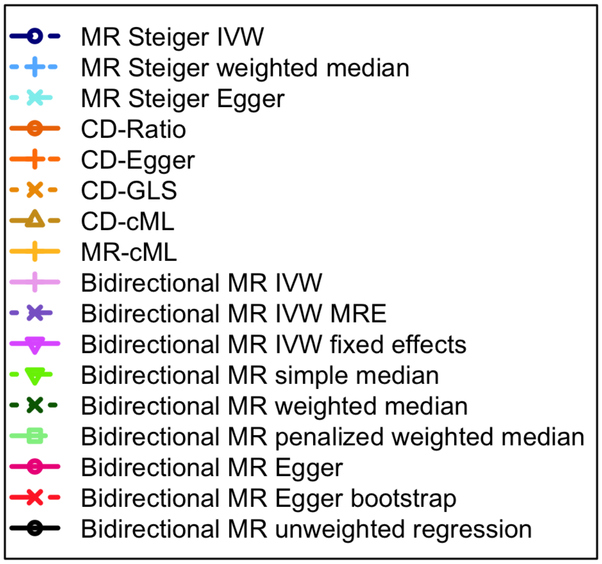

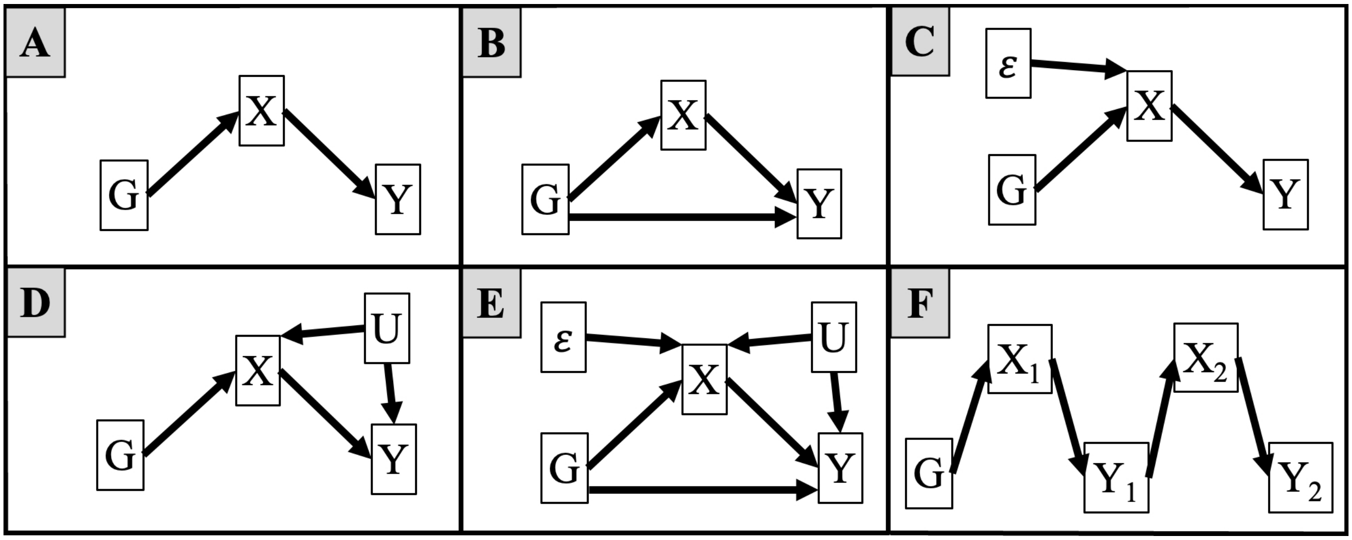

### 1 Phenotype 1: normally distributed; Phenotype 2: normally distributed; sample size $N = 1{,}000$; SNP effect size of 1.

**Supplemental Table 1.** Simulation results with $N = 1{,}000$ and 10 SNPs ($\gamma_{GX} = 1$, $\beta_{GY} = 1$). Heat maps summarize performance across the range of $\beta_X$ values shown in Supplemental Figure 1. Green cells indicate consistently high performance across all checks (no violations detected) in at least one setting. Otherwise, overlaid stripes show specific violations: inflated type I error ($> 10\%$, red), spurious $Y \to X$ inference ($> 5\%$, orange), or low power to detect $X \to Y$ ($< 75\%$, blue).

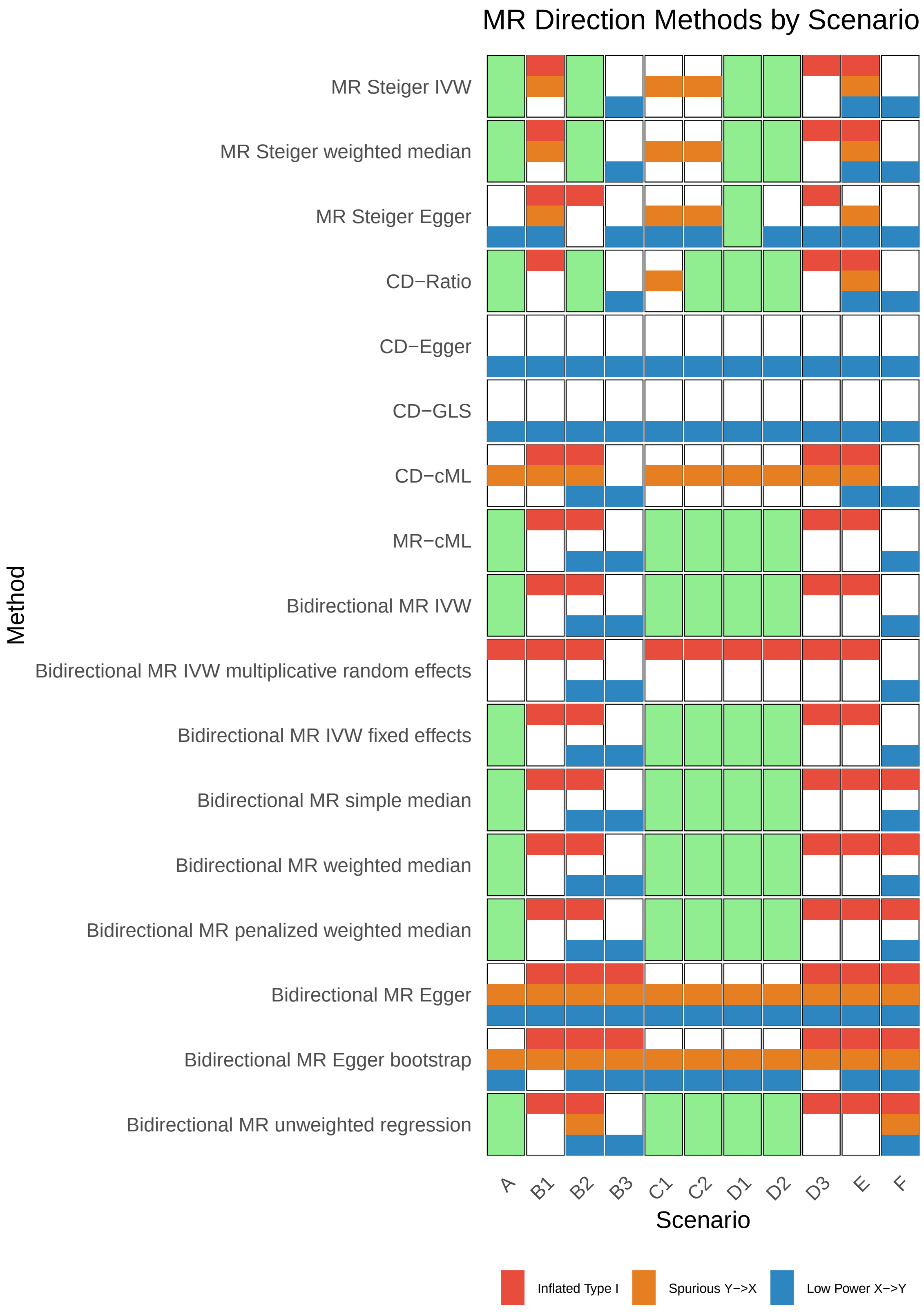

**Supplemental Figure 1.** Simulation results for Scenario 1 ($N = 1{,}000$, 10 SNPs, $\gamma_{GX} = 1$, $\beta_{GY} = 1$). Phenotypes 1 and 2 are continuous (normally distributed). Columns correspond to sub-scenarios (A–F); rows show the proportion of simulations returning case 1 ($X \rightarrow Y$, top) or case 2 ($Y \rightarrow X$, bottom) across values of $\beta_X$. The $x$-axis shows the true effect size $\beta_X$, and the $y$-axis shows the proportion of simulations where each method returned the given case. Line colors correspond to MR methods (see overview legend).

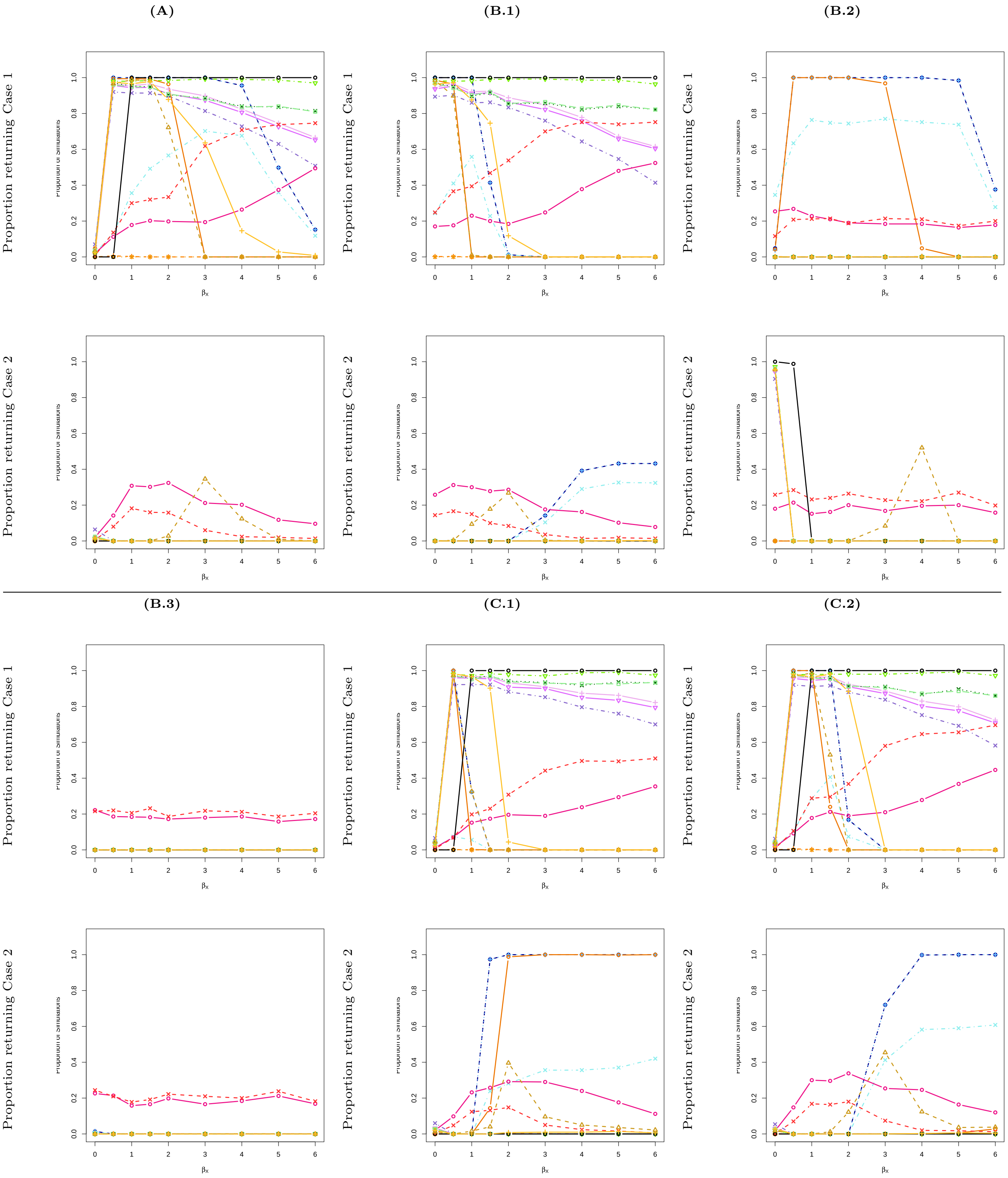

**(D.1)**

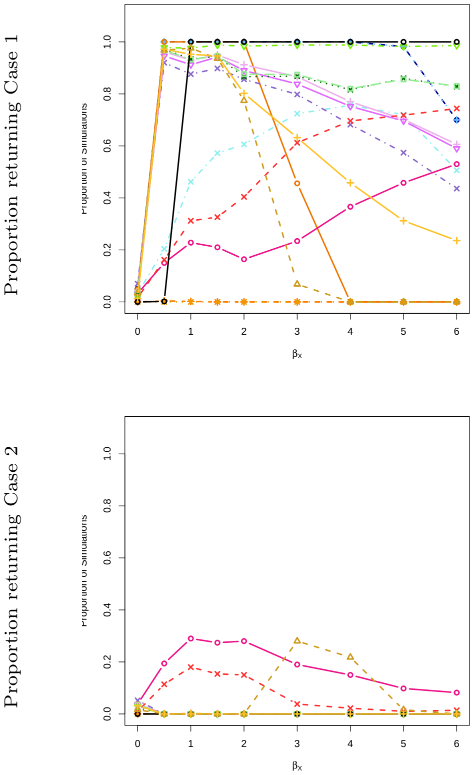
Proportion returning Case 1
Proportion returning Case 2
0.0 0.2 0.4 0.6 0.8 1.0
0 1 2 3 4 5 6
βx

**(D.2)**

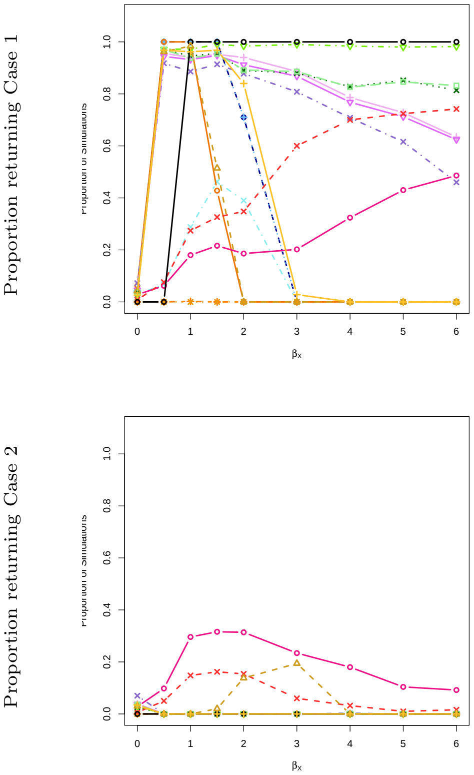
Proportion returning Case 1
Proportion returning Case 2
0.0 0.2 0.4 0.6 0.8 1.0
0 1 2 3 4 5 6
βx

**(D.3)**

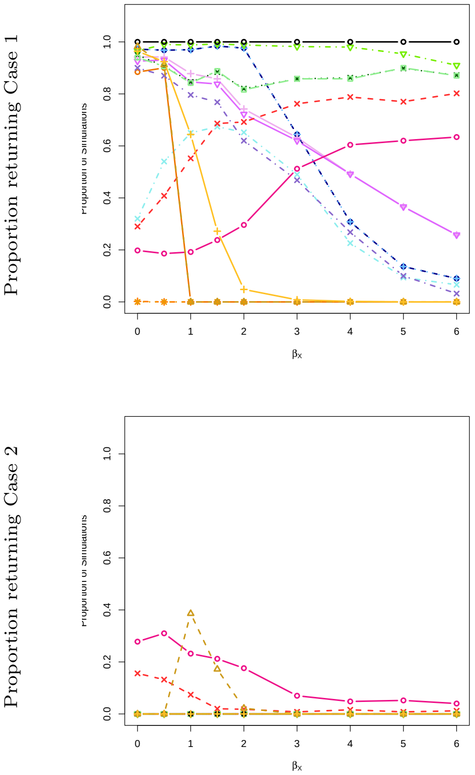
Proportion returning Case 1
Proportion returning Case 2
0.0 0.2 0.4 0.6 0.8 1.0
0 1 2 3 4 5 6
βx

**(E)**

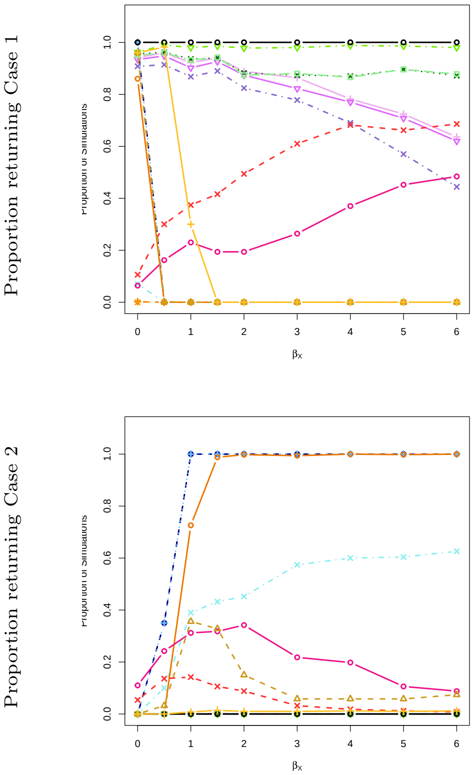
Proportion returning Case 1
Proportion returning Case 2
0.0 0.2 0.4 0.6 0.8 1.0
0 1 2 3 4 5 6
βx

**(F)**

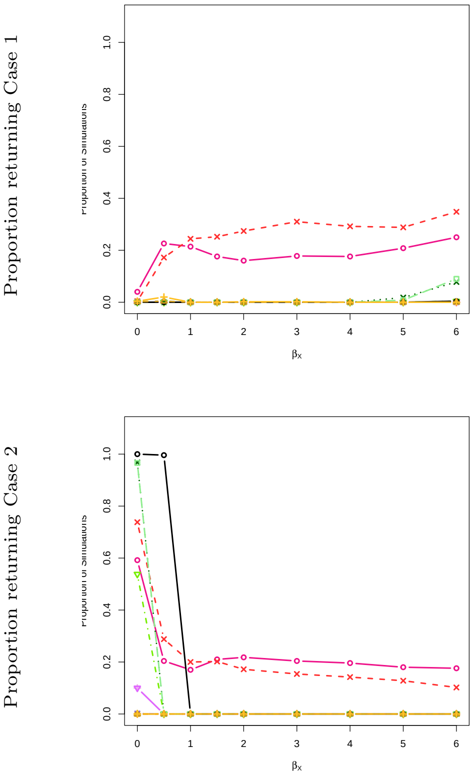
Proportion returning Case 1
Proportion returning Case 2
0.0 0.2 0.4 0.6 0.8 1.0
0 1 2 3 4 5 6
βx

#### 2 Phenotype 1: Bernoulli distributed; Phenotype 2: normally distributed; sample size $N = 1{,}000$; SNP effect size of 1.

**Supplemental Table 2.** Simulation results with $N = 1{,}000$ and 10 SNPs ($\gamma_{GX} = 1$, $\beta_{GY} = 1$). Heat maps summarize performance across the range of $\beta_X$ values shown in Supplemental Figure 2. Green cells indicate consistently high performance across all checks (no violations detected) in at least one setting. Otherwise, overlaid stripes show specific violations: inflated type I error ($> 10\%$, red), spurious $Y \to X$ inference ($> 5\%$, orange), or low power to detect $X \to Y$ ($< 75\%$, blue).

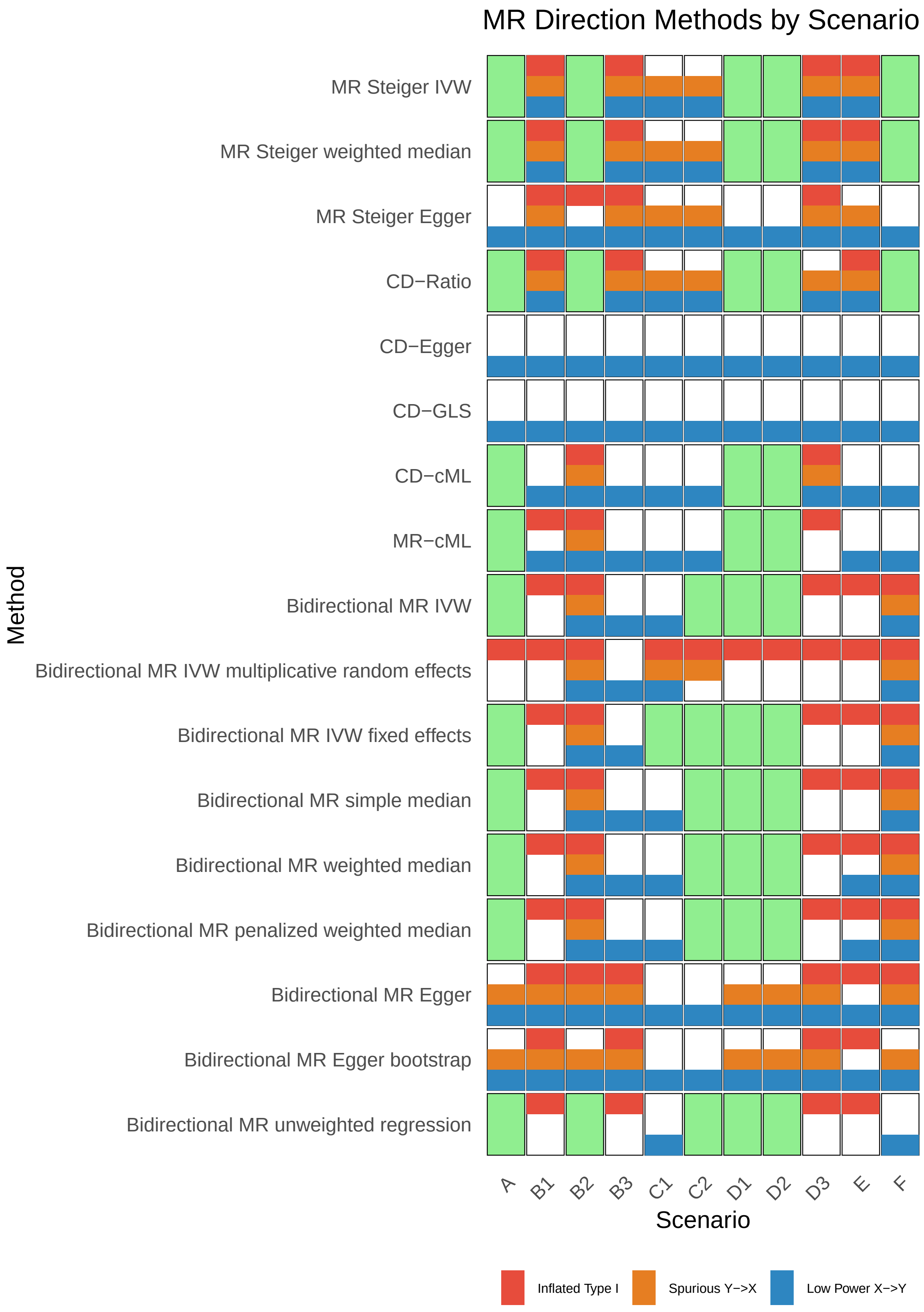

**Supplemental Figure 2.** Simulation results for Scenario 2 ($N$ = 1,000, 10 SNPs, $\gamma_{GX}$ = 1, $\beta_{GY}$ = 1). Phenotype 1 is binary (Bernoulli distributed) and phenotype 2 is continuous (normally distributed). Columns correspond to sub-scenarios (A–F); rows show the proportion of simulations returning case 1 ($X \rightarrow Y$, top) or case 2 ($Y \rightarrow X$, bottom) across values of $\beta_X$. The $x$-axis shows the true effect size $\beta_X$, and the $y$-axis shows the proportion of simulations where each method returned the given case. Line colors correspond to MR methods (see overview legend).

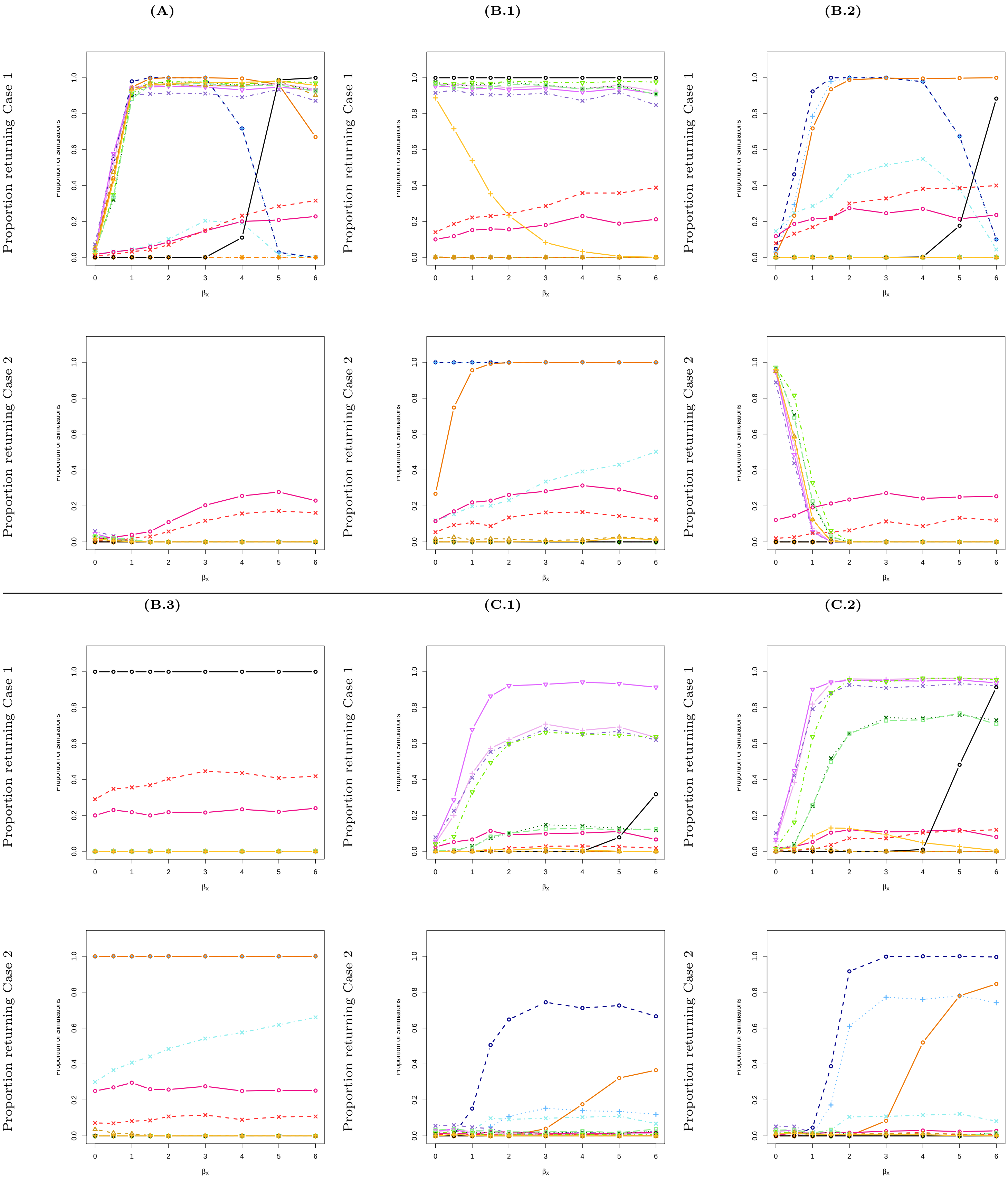

**(D.1)**

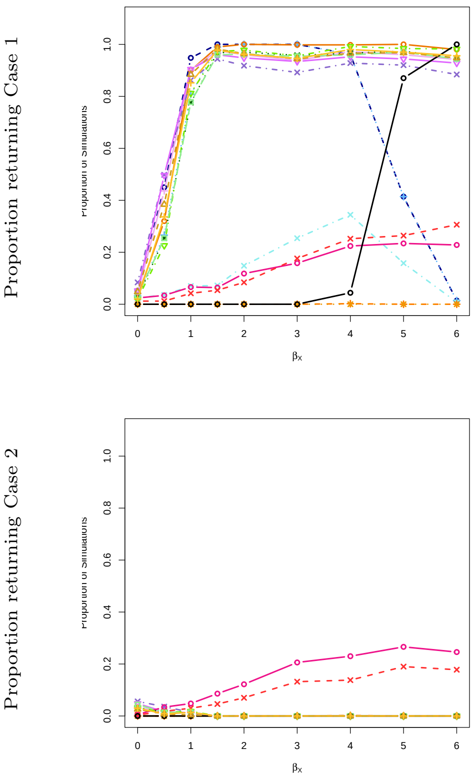
Proportion returning Case 1
Proportion returning Case 2
$\beta_x$

**(D.2)**

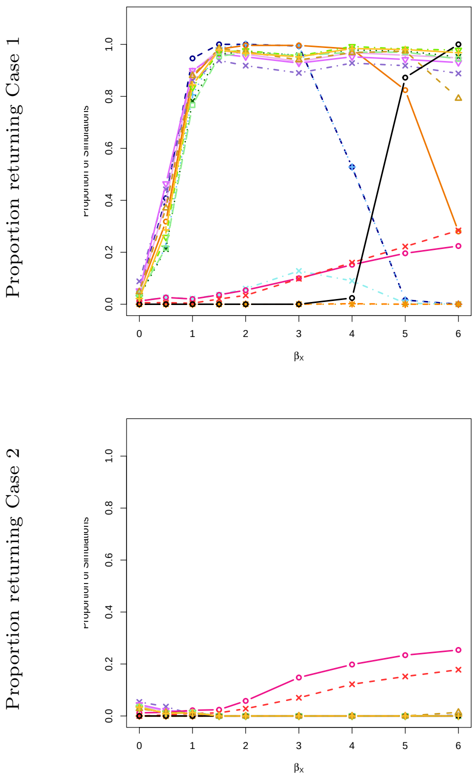
Proportion returning Case 1
Proportion returning Case 2
$\beta_x$

**(D.3)**

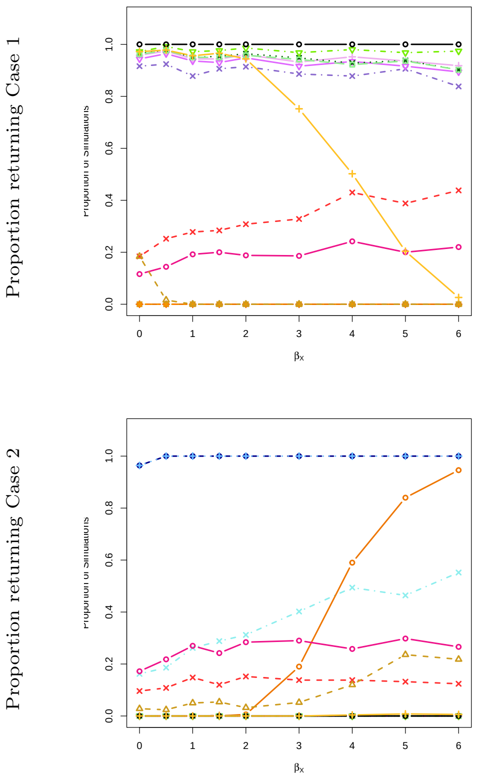
Proportion returning Case 1
Proportion returning Case 2
$\beta_x$

**(E)**

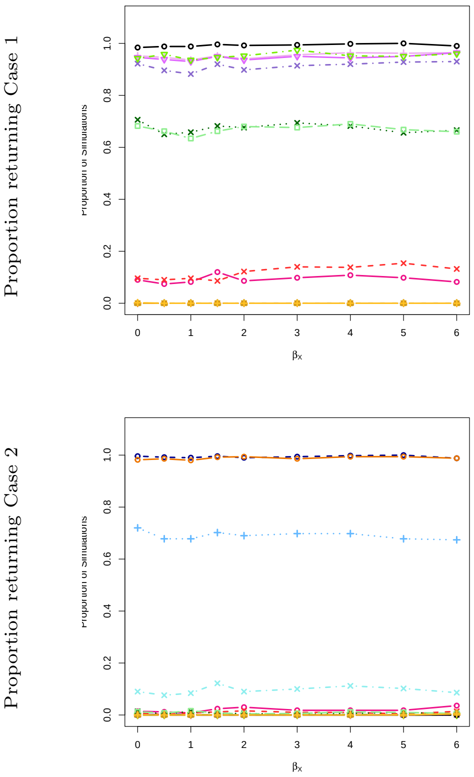
Proportion returning Case 1
Proportion returning Case 2
$\beta_x$

**(F)**

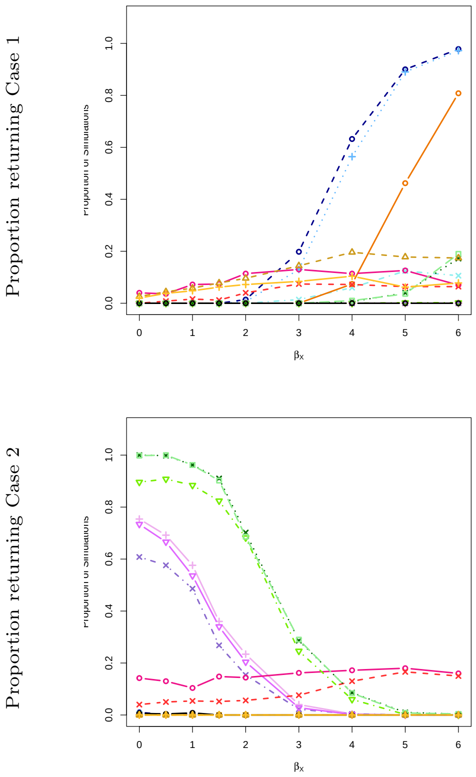
Proportion returning Case 1
Proportion returning Case 2
$\beta_x$

### 3 Phenotype 1: normally distributed; Phenotype 2: Bernoulli distributed; sample size $N = 1{,}000$; SNP effect size of 1.

**Supplemental Table 3.** Simulation results with $N = 1{,}000$ and 10 SNPs ($\gamma_{GX} = 1$, $\beta_{GY} = 1$). Heat maps summarize performance across the range of $\beta_X$ values shown in Supplemental Figure 3. Green cells indicate consistently high performance across all checks (no violations detected) in at least one setting. Otherwise, overlaid stripes show specific violations: inflated type I error ($> 10\%$, red), spurious $Y \to X$ inference ($> 5\%$, orange), or low power to detect $X \to Y$ ($< 75\%$, blue).

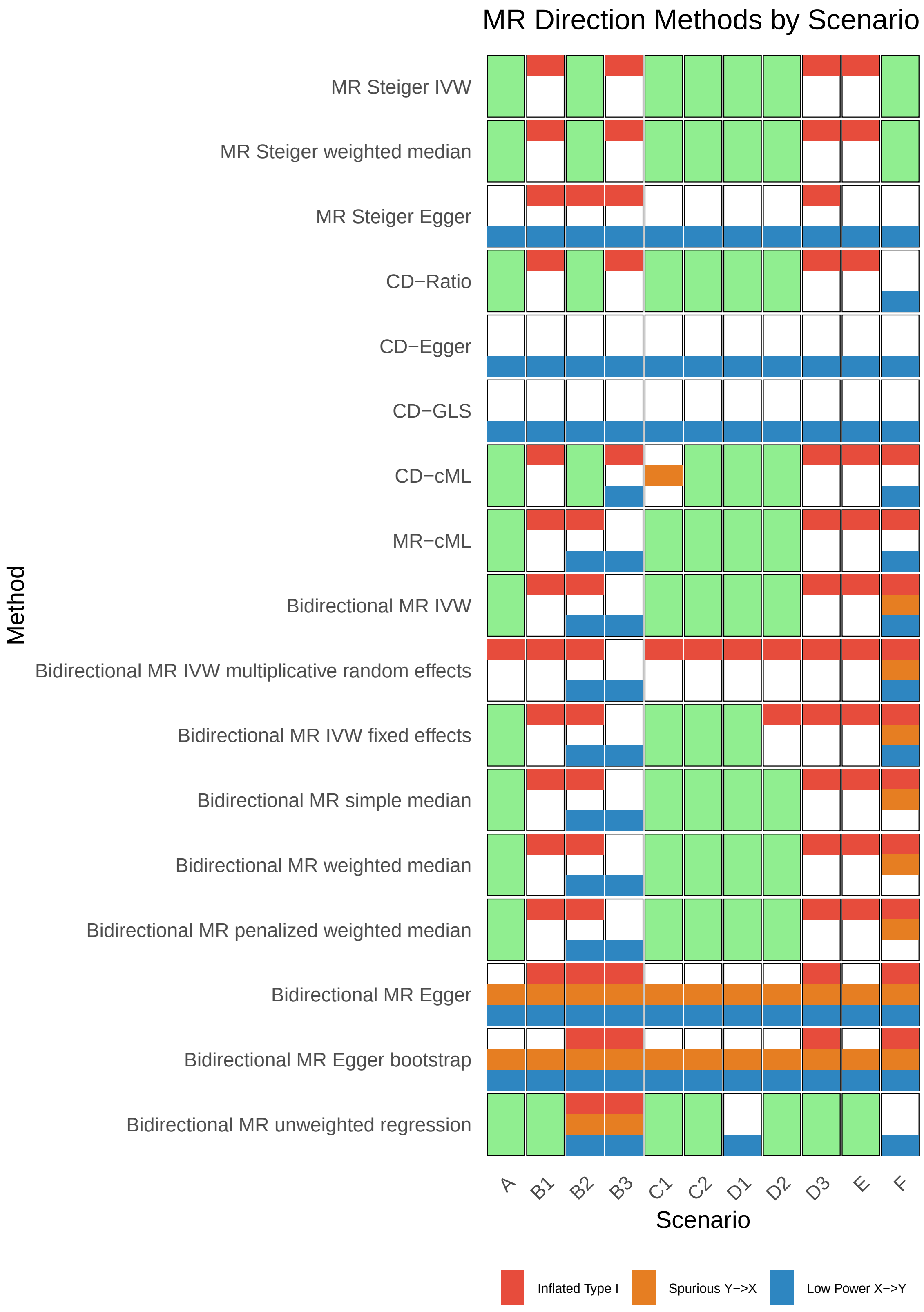

**Supplemental Figure 3.** Simulation results for Scenario 3 ($N$ = 1,000, 10 SNPs, $\gamma_{GX} = 1$, $\beta_{GY} = 1$). Phenotype 1 is continuous (normally distributed) and phenotype 2 is binary (Bernoulli distributed). Columns correspond to sub-scenarios (A–F); rows show the proportion of simulations returning case 1 ($X \to Y$, top) or case 2 ($Y \to X$, bottom) across values of $\beta_X$. The $x$-axis shows the true effect size $\beta_X$, and the $y$-axis shows the proportion of simulations where each method returned the given case. Line colors correspond to MR methods (see overview legend).

**(A)**

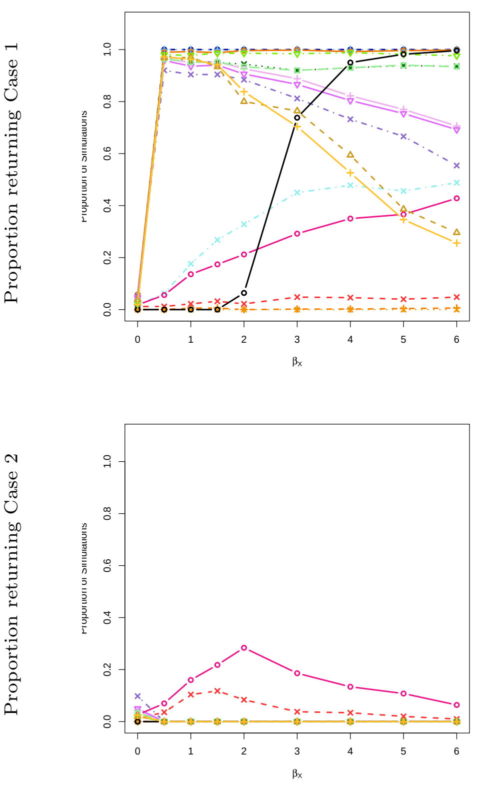

**(B.1)**

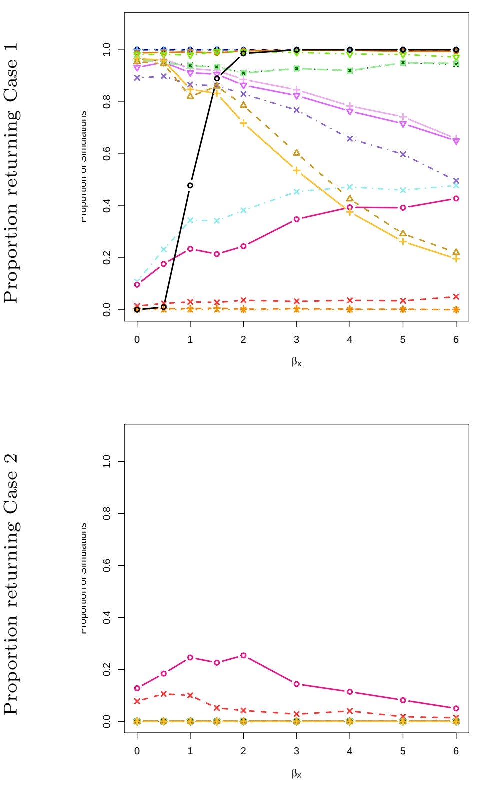

**(B.2)**

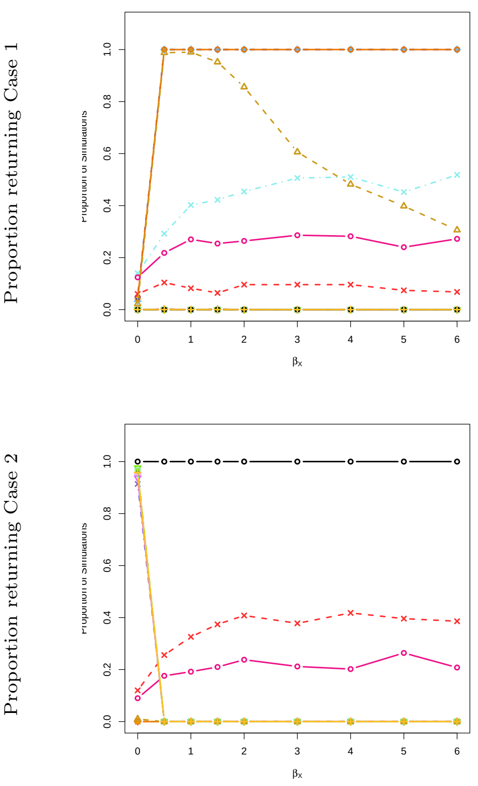

**(B.3)**

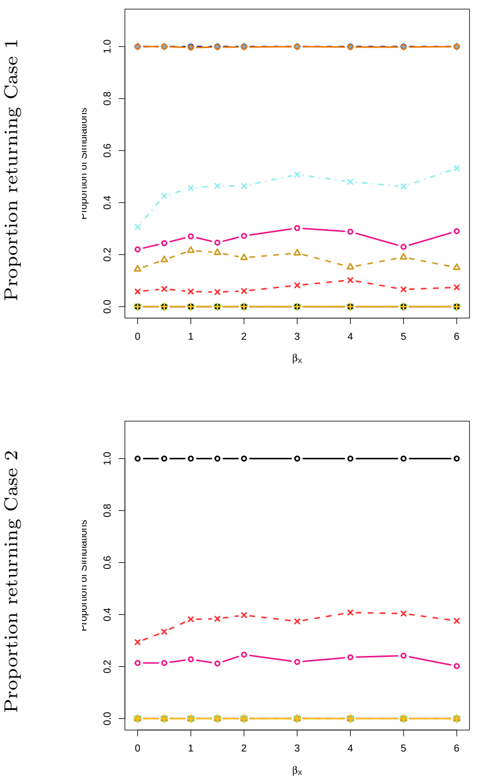

**(C.1)**

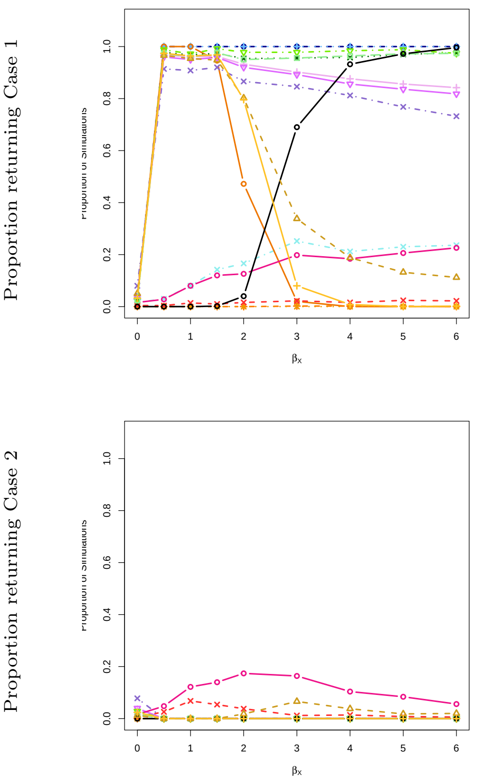

**(C.2)**

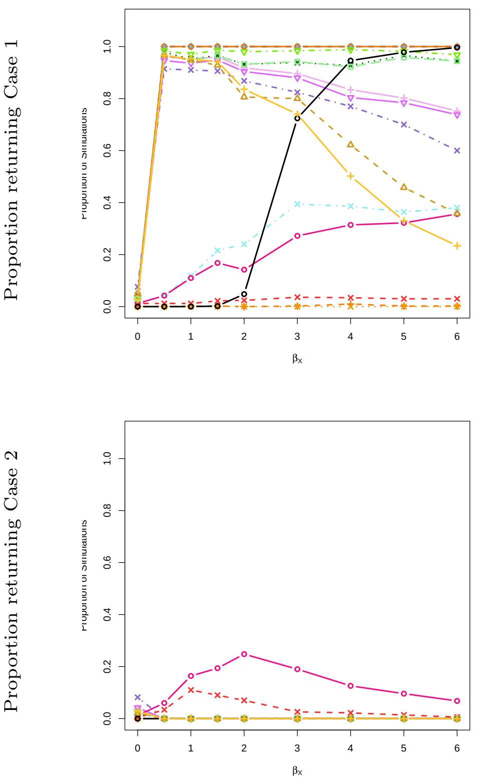

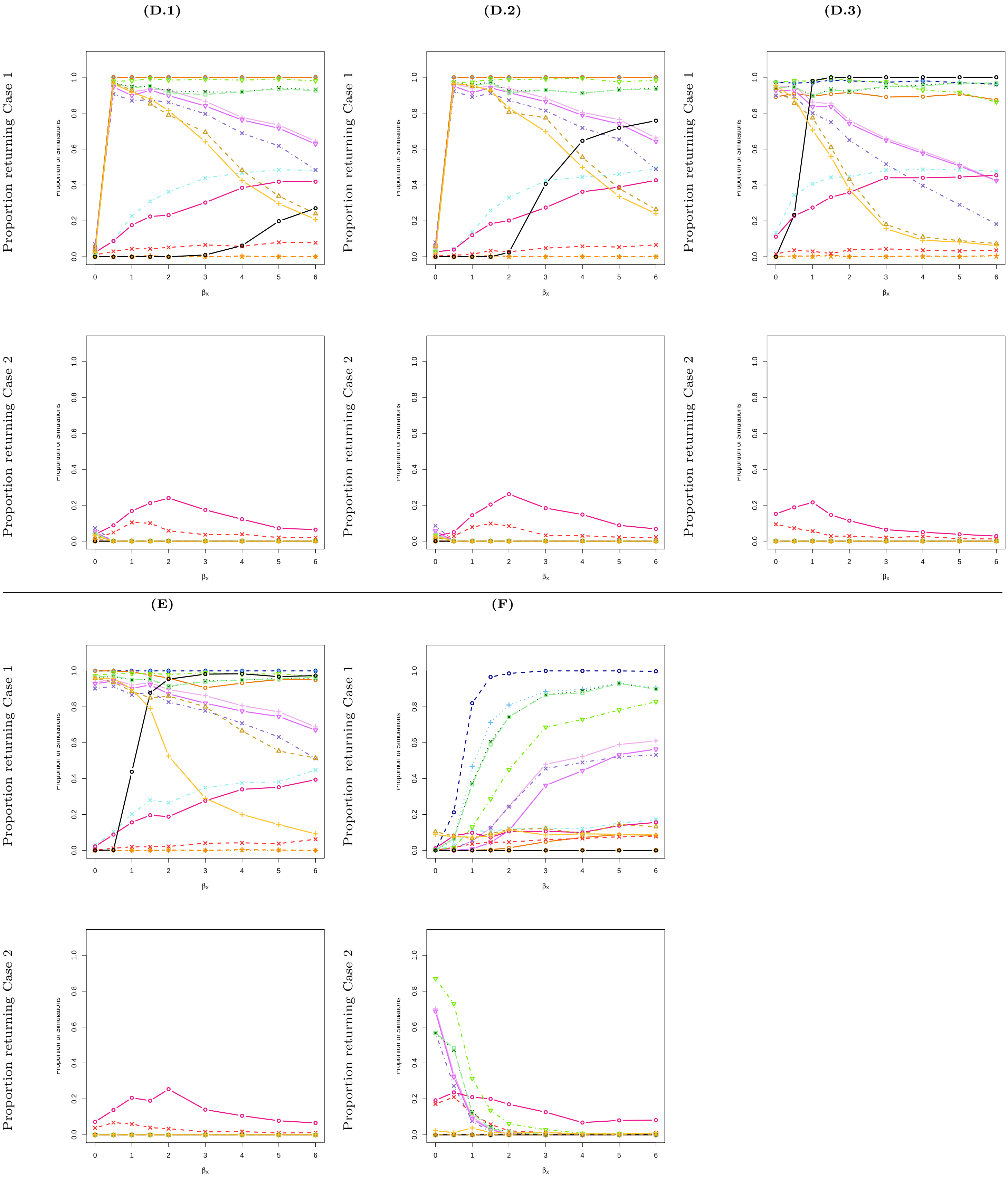
(D.1)
(D.2)
(D.3)
(E)
(F)
Proportion returning Case 1
Proportion returning Case 2
Proportion of simulations
0.0
0.2
0.4
0.6
0.8
1.0
0
1
2
3
4
5
6
βx

#### 4 Phenotype 1: Bernoulli distributed; Phenotype 2: Bernoulli distributed; sample size $N = 1{,}000$; SNP effect size of 1.

**Supplemental Table 4.** Simulation results with $N = 1{,}000$ and 10 SNPs ($\gamma_{GX} = 1$, $\beta_{GY} = 1$). Heat maps summarize performance across the range of $\beta_X$ values shown in Supplemental Figure 4. Green cells indicate consistently high performance across all checks (no violations detected) in at least one setting. Otherwise, overlaid stripes show specific violations: inflated type I error (> 10%, red), spurious $Y \to X$ inference (> 5%, orange), or low power to detect $X \to Y$ (< 75%, blue).

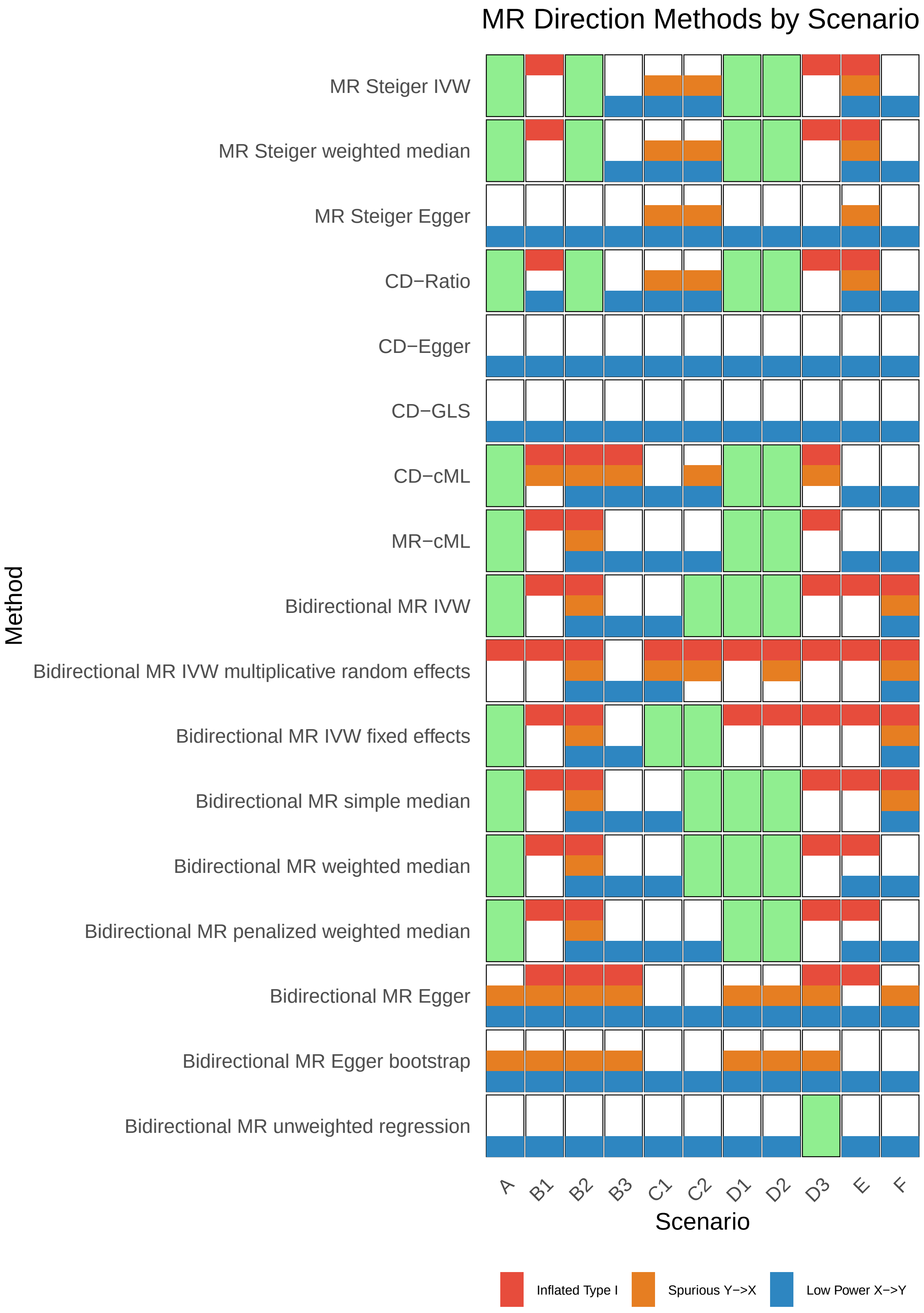

**Supplemental Figure 4.** Simulation results for Scenario 4 ($N = 1{,}000$, 10 SNPs, $\gamma_{GX} = 1$, $\beta_{GY} = 1$). Both phenotypes are binary (Bernoulli distributed). Columns correspond to sub-scenarios (A–F); rows show the proportion of simulations returning case 1 ($X \rightarrow Y$, top) or case 2 ($Y \rightarrow X$, bottom) across values of $\beta_X$. The $x$-axis shows the true effect size $\beta_X$, and the $y$-axis shows the proportion of simulations where each method returned the given case. Line colors correspond to MR methods (see overview legend).

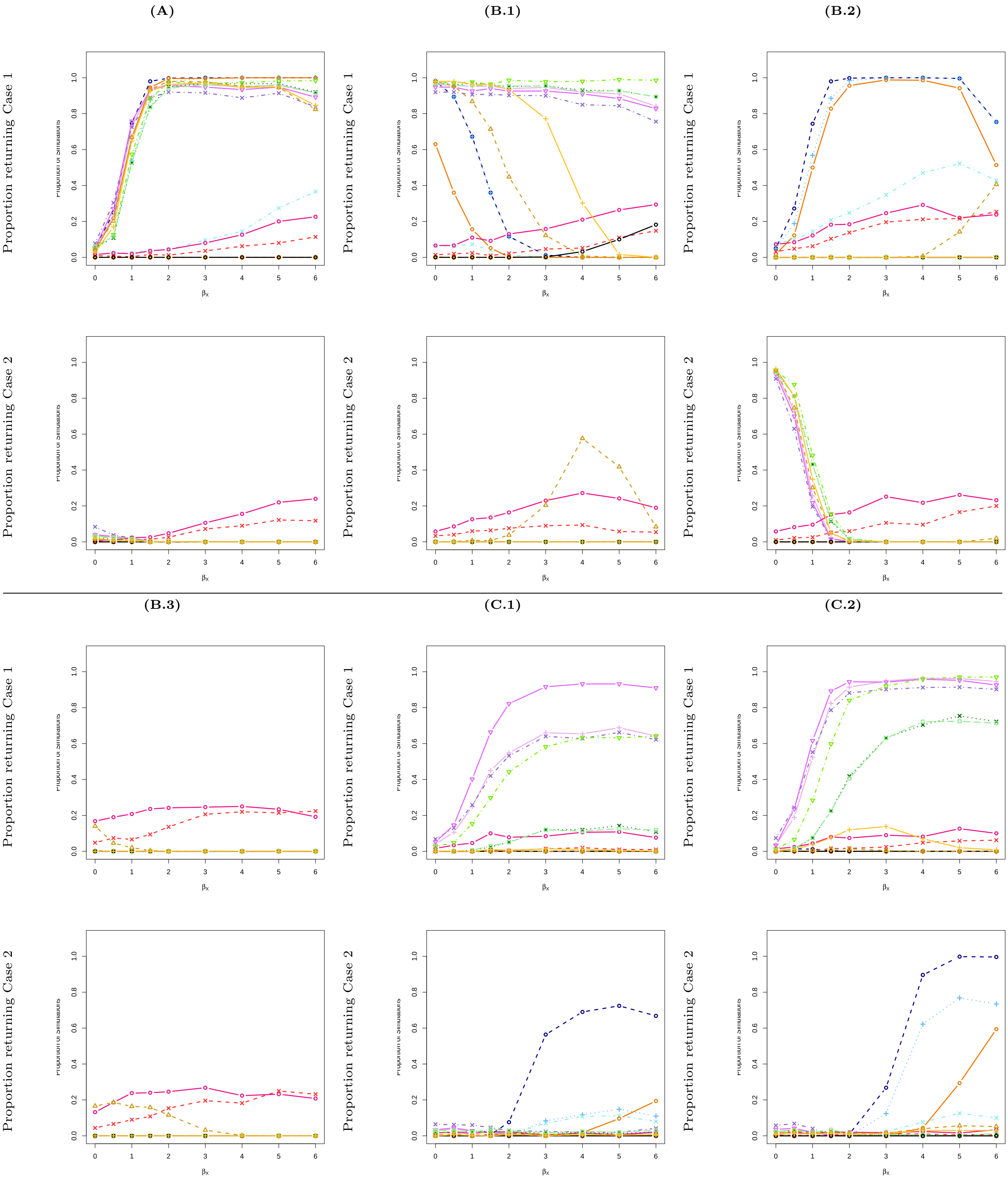

**(D.1)** **(D.2)** **(D.3)**

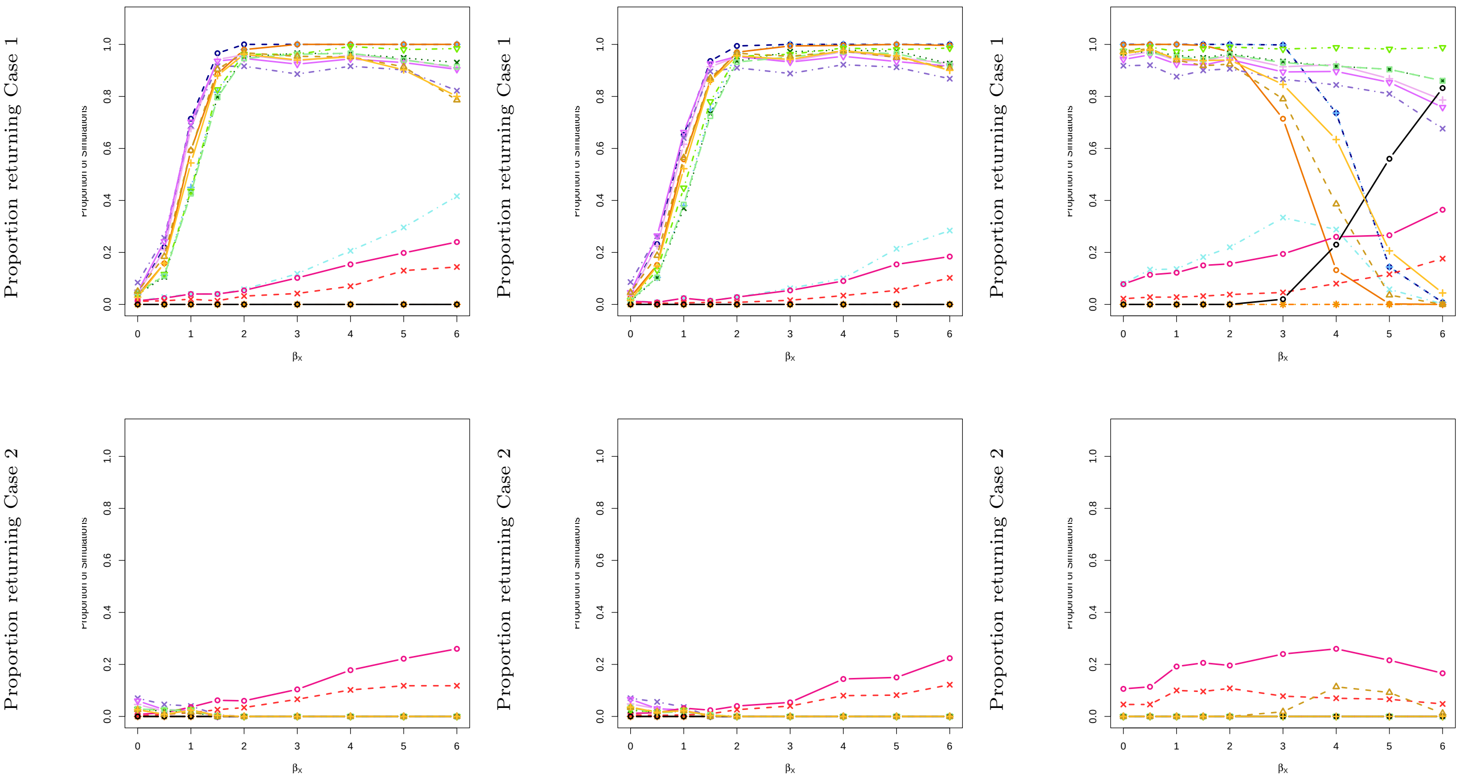
Proportion returning Case 1
Proportion returning Case 2
βx

**(E)** **(F)**

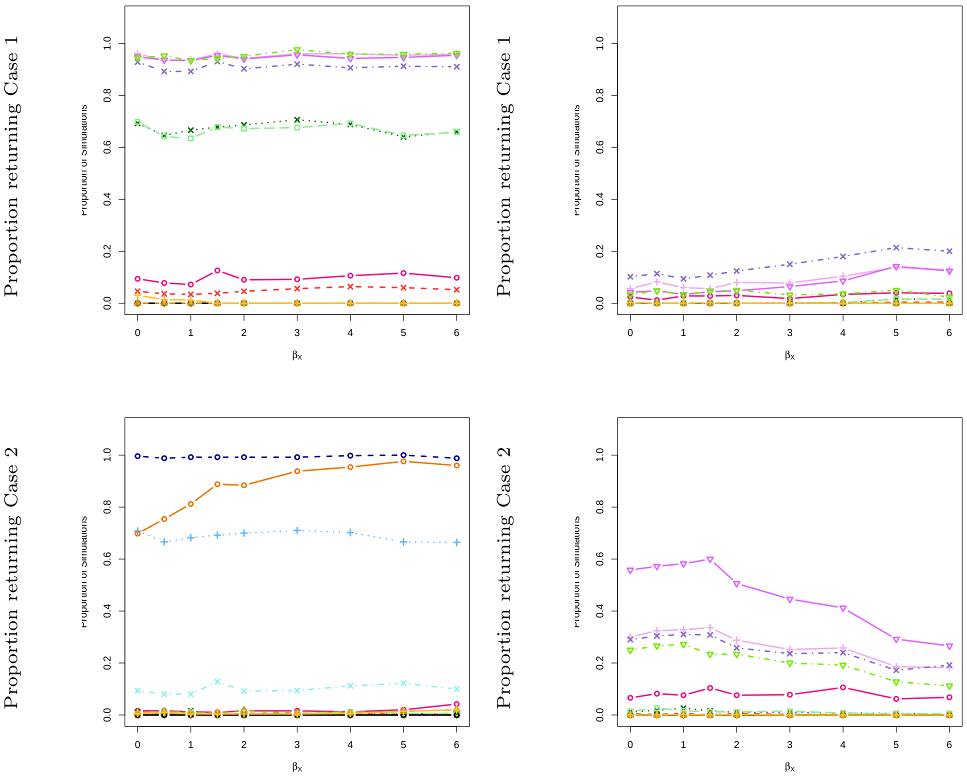
Proportion returning Case 1
Proportion returning Case 2
βx

### 5 Phenotype 1: Bernoulli distributed; Phenotype 2: normally distributed; sample size $N = 100{,}000$; SNP effect size of 0.2.

**Supplemental Table 5.** Simulation results with $N = 100{,}000$ and 50 SNPs ($\gamma_{GX} = 0.2$, $\beta_{GY} = 0.2$). Heat maps summarize performance across the range of $\beta_X$ values shown in Supplemental Figure 5. Green cells indicate consistently high performance across all checks (no violations detected) in at least one setting. Otherwise, overlaid stripes show specific violations: inflated type I error ($> 10\%$, red), spurious $Y \to X$ inference ($> 5\%$, orange), or low power to detect $X \to Y$ ($< 75\%$, blue).

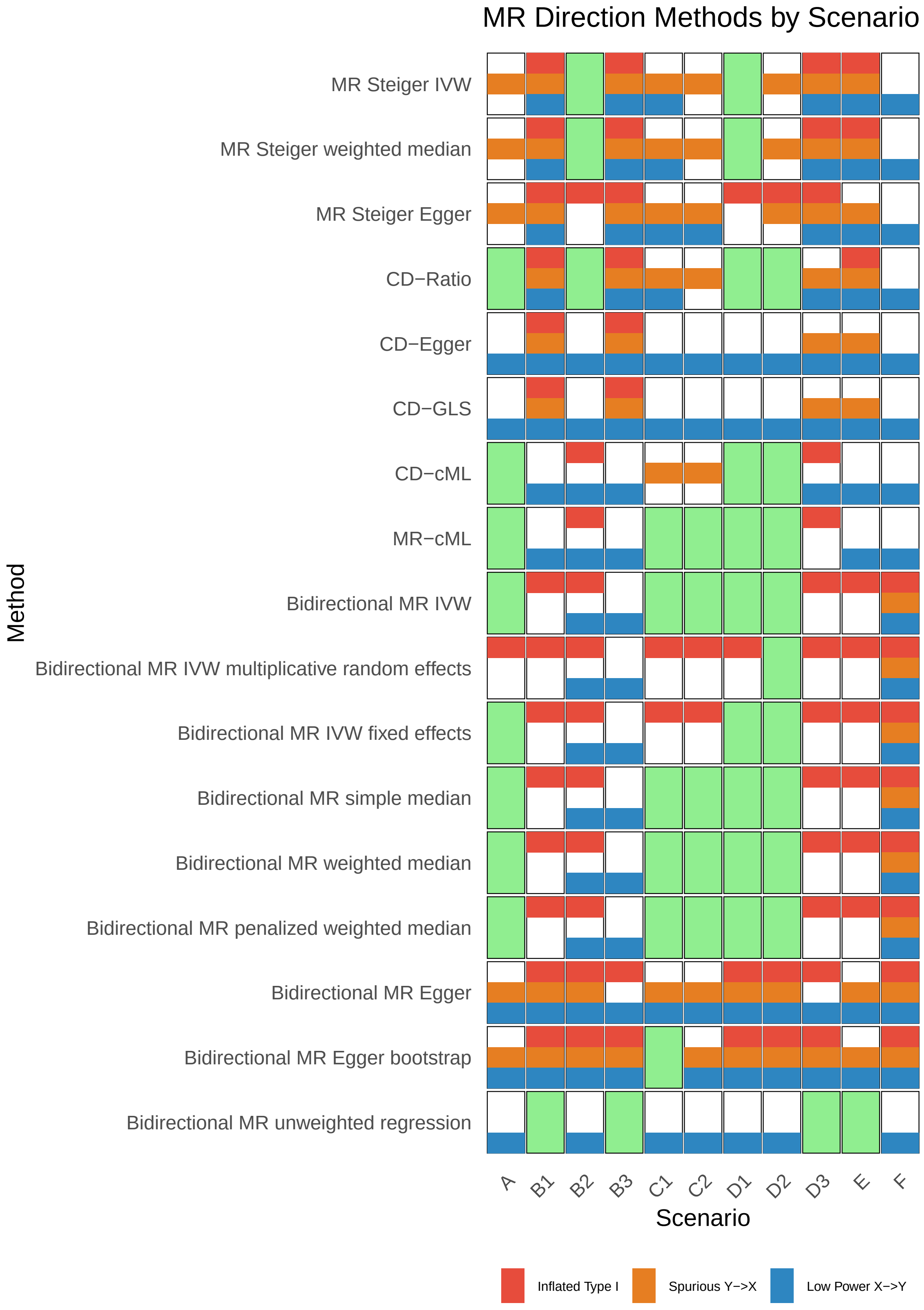

**Supplemental Figure 5.** Simulation results for Scenario 6 ($N$ = 100,000, 50 SNPs, $\gamma_{GX} = 0.2$, $\beta_{GY} = 0.2$). Phenotype 1 is binary (Bernoulli distributed) and phenotype 2 is continuous (normally distributed). Columns correspond to sub-scenarios (A–F); rows show the proportion of simulations returning case 1 ($X \to Y$, top) or case 2 ($Y \to X$, bottom) across values of $\beta_X$. The $x$-axis shows the true effect size $\beta_X$, and the $y$-axis shows the proportion of simulations where each method returned the given case. Line colors correspond to MR methods (see overview legend).

**(A)**

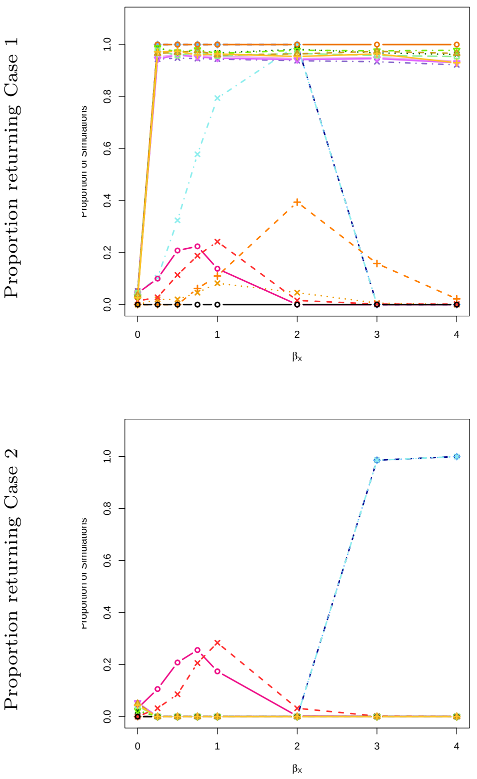

**(B.1)**

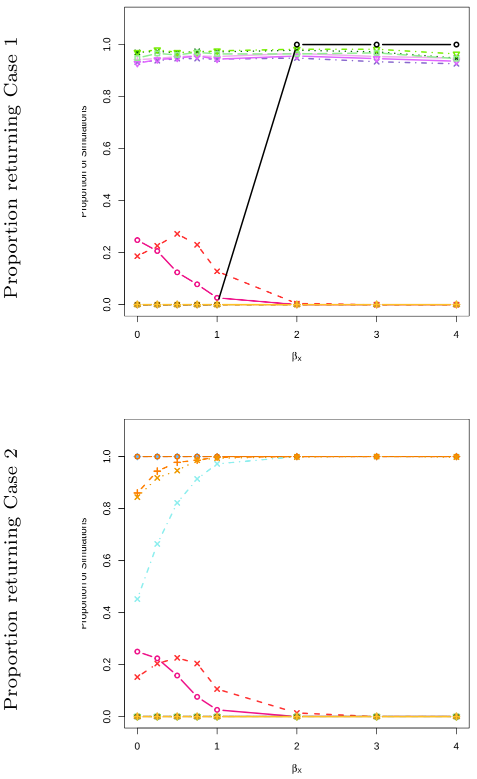

**(B.2)**

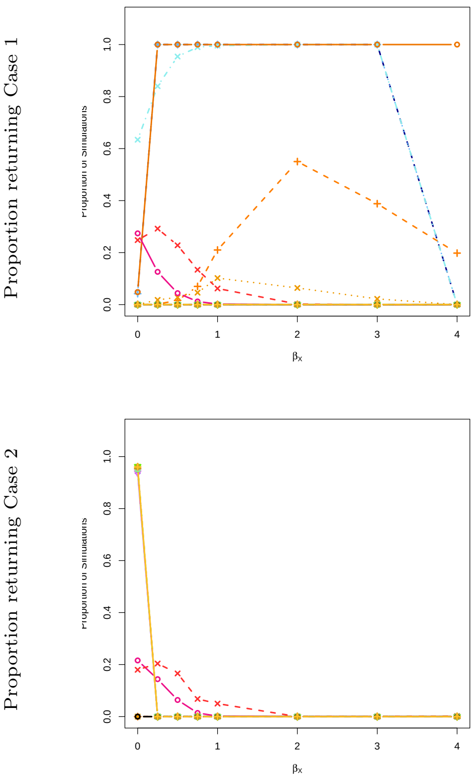

**(B.3)**

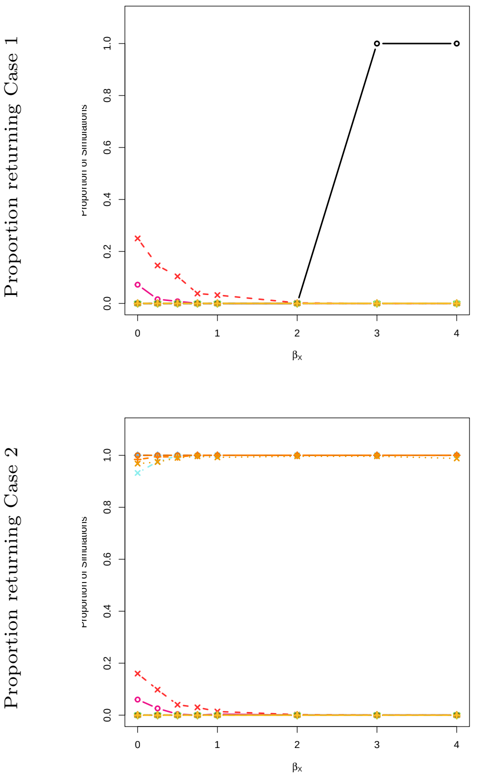

**(C.1)**

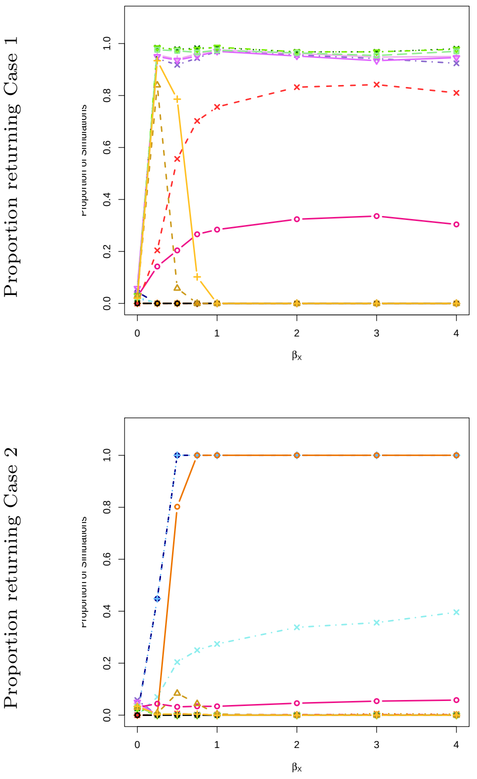

**(C.2)**

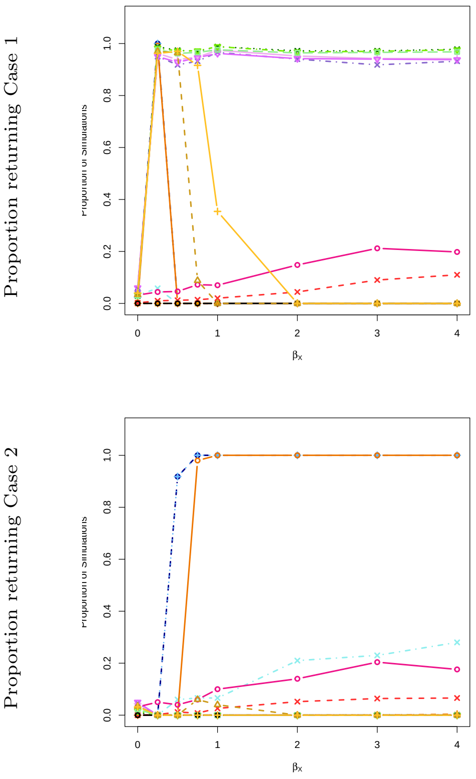

**(D.1)**

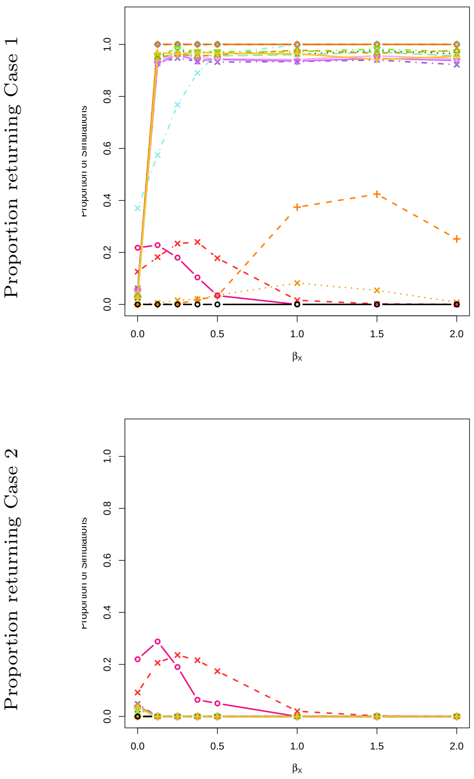
Proportion returning Case 1
Proportion returning Case 2
$\beta_x$

**(D.2)**

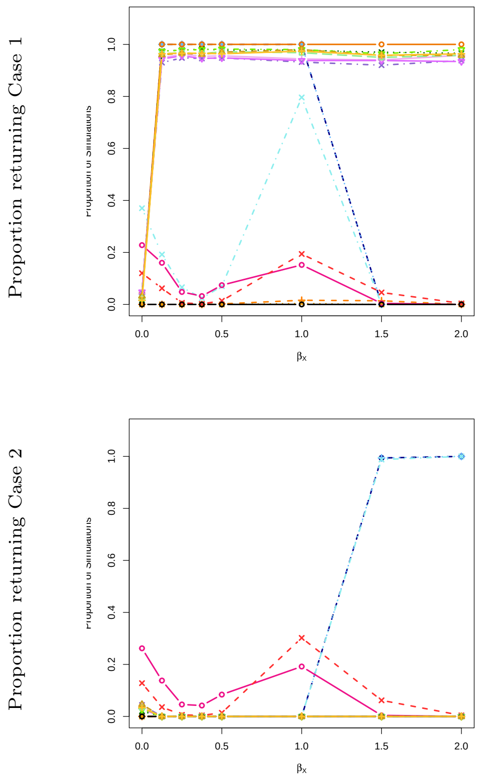
Proportion returning Case 1
Proportion returning Case 2
$\beta_x$

**(D.3)**

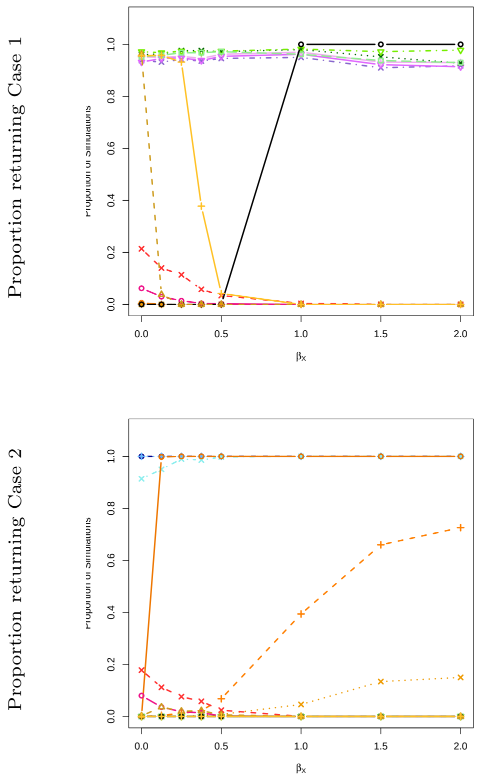
Proportion returning Case 1
Proportion returning Case 2
$\beta_x$

**(E)**

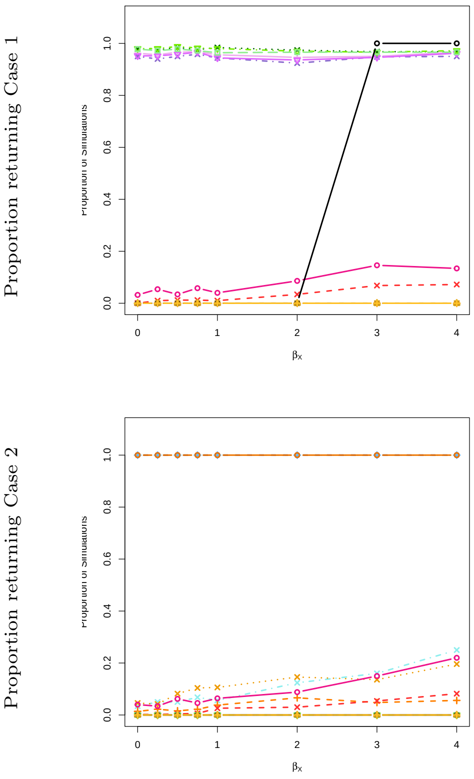
Proportion returning Case 1
Proportion returning Case 2
$\beta_x$

**(F)**

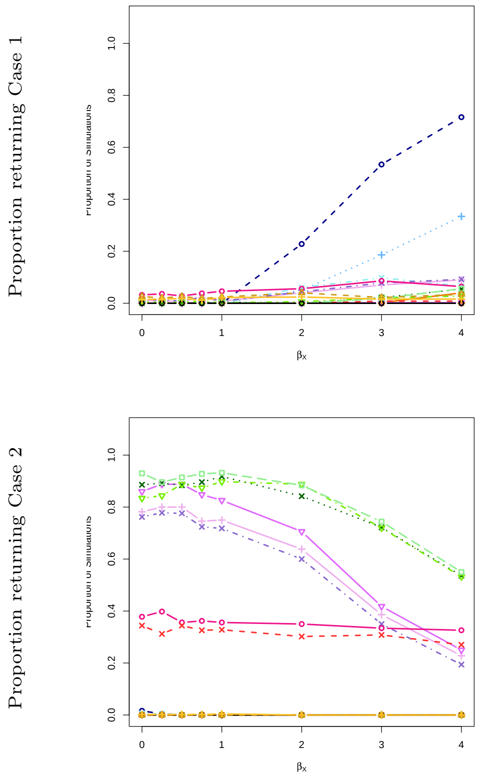
Proportion returning Case 1
Proportion returning Case 2
$\beta_x$

### 6 Phenotype 1: normally distributed; Phenotype 2: Bernoulli distributed; sample size $N = 100{,}000$; SNP effect size of 0.2.

**Supplemental Table 6.** Simulation results with $N = 100{,}000$ and 50 SNPs ($\gamma_{GX} = 0.2$, $\beta_{GY} = 0.2$). Heat maps summarize performance across the range of $\beta_X$ values shown in Supplemental Figure 6. Green cells indicate consistently high performance across all checks (no violations detected) in at least one setting. Otherwise, overlaid stripes show specific violations: inflated type I error ($> 10\%$, red), spurious $Y \rightarrow X$ inference ($> 5\%$, orange), or low power to detect $X \rightarrow Y$ ($< 75\%$, blue).

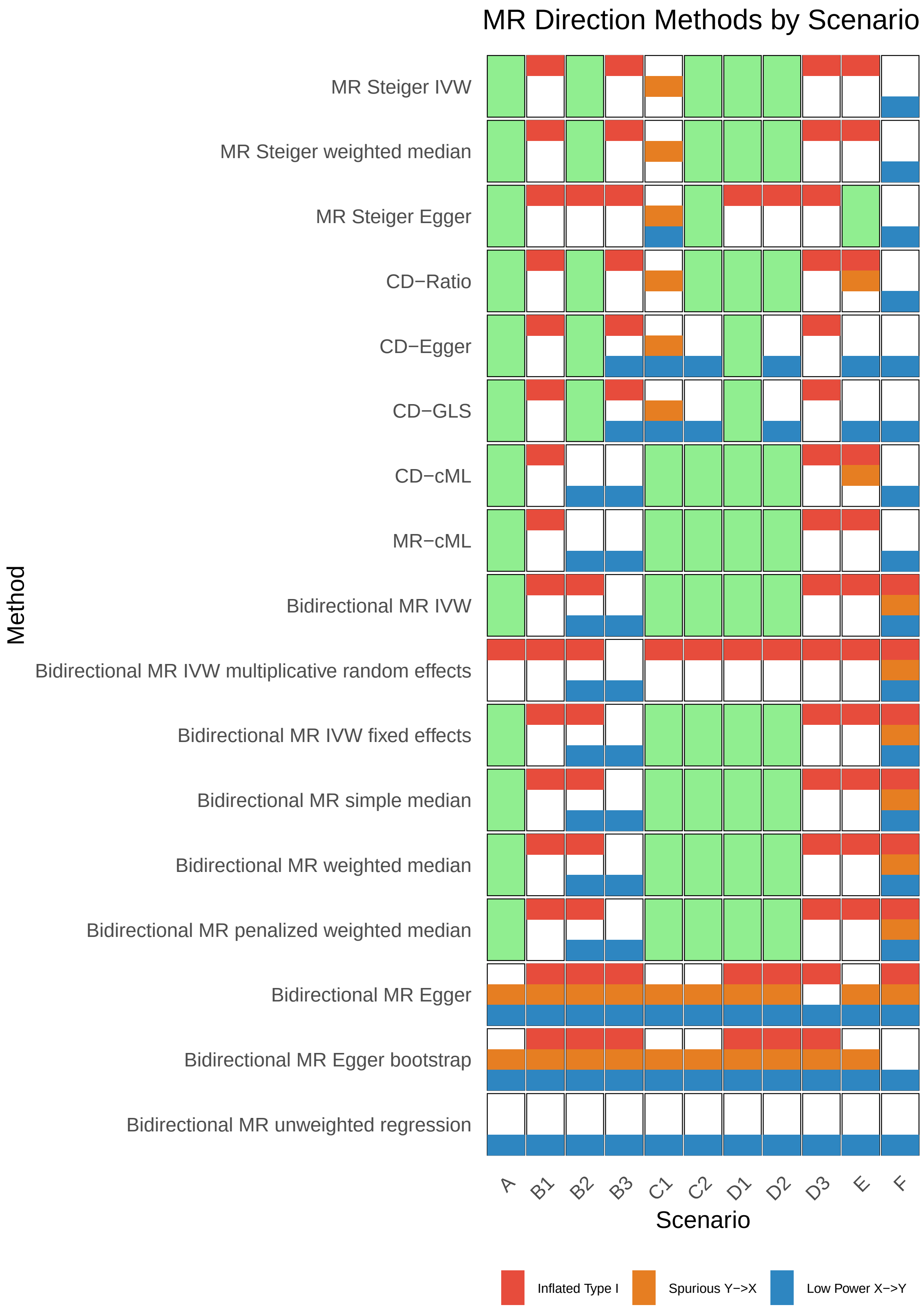

**Supplemental Figure 6.** Simulation results for Scenario 7 ($N$ = 100,000, 50 SNPs, $\gamma_{GX} = 0.2$, $\beta_{GY} = 0.2$). Phenotype 1 is continuous (normally distributed) and phenotype 2 is binary (Bernoulli distributed). Columns correspond to sub-scenarios (A–F); rows show the proportion of simulations returning case 1 ($X \to Y$, top) or case 2 ($Y \to X$, bottom) across values of $\beta_X$. The $x$-axis shows the true effect size $\beta_X$, and the $y$-axis shows the proportion of simulations where each method returned the given case. Line colors correspond to MR methods (see overview legend).

**(A)**

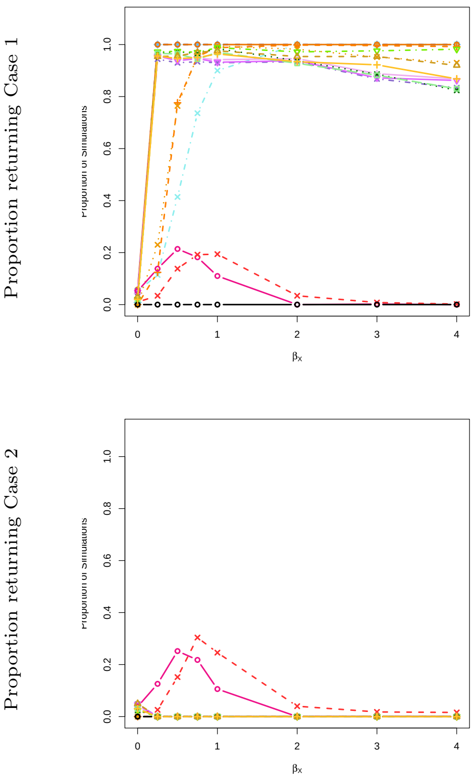

**(B.1)**

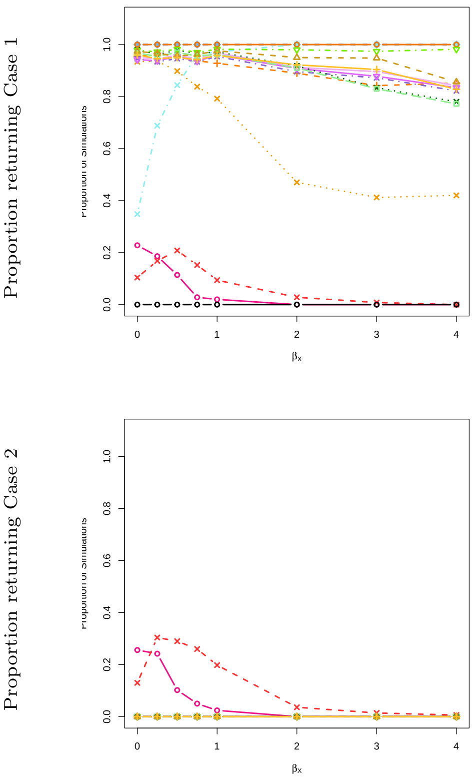

**(B.2)**

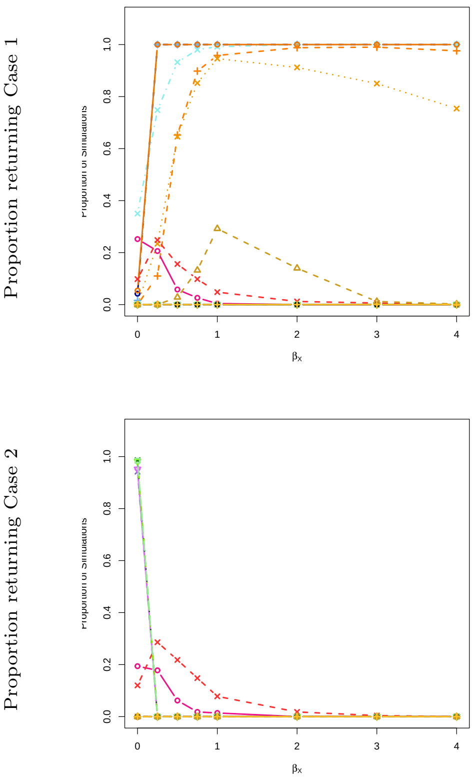

**(B.3)**

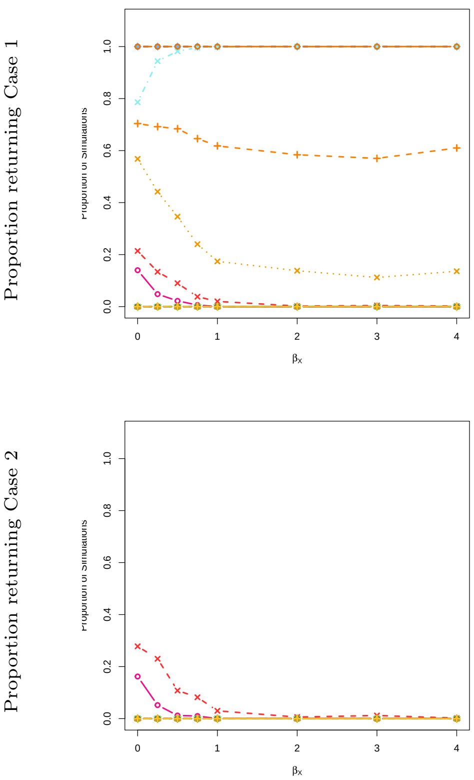

**(C.1)**

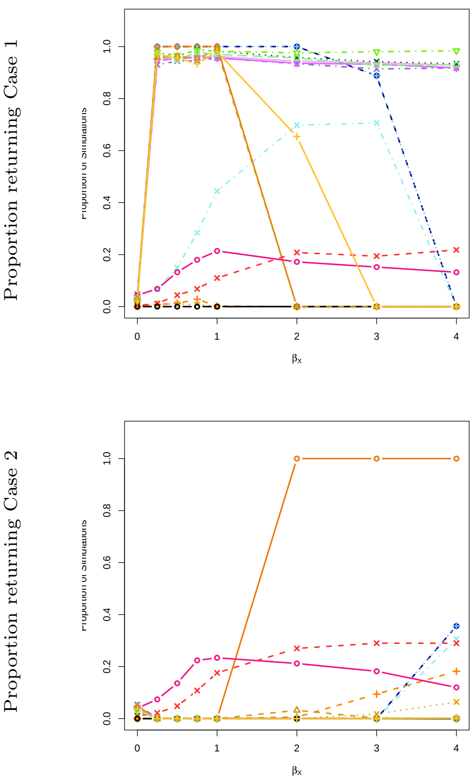

**(C.2)**

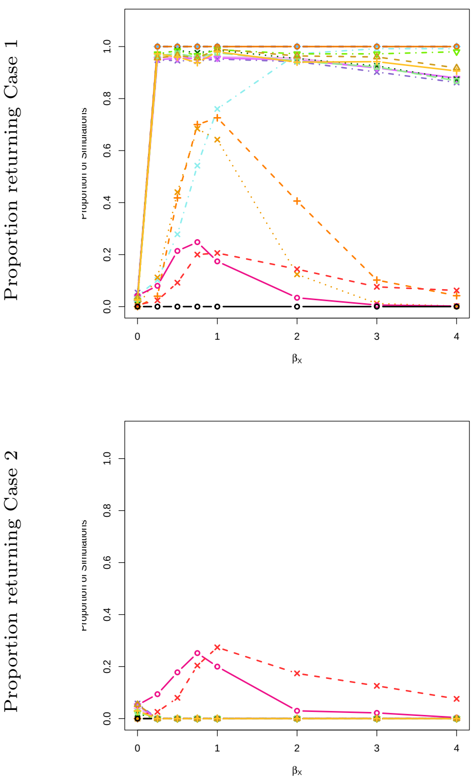

**(D.1)**

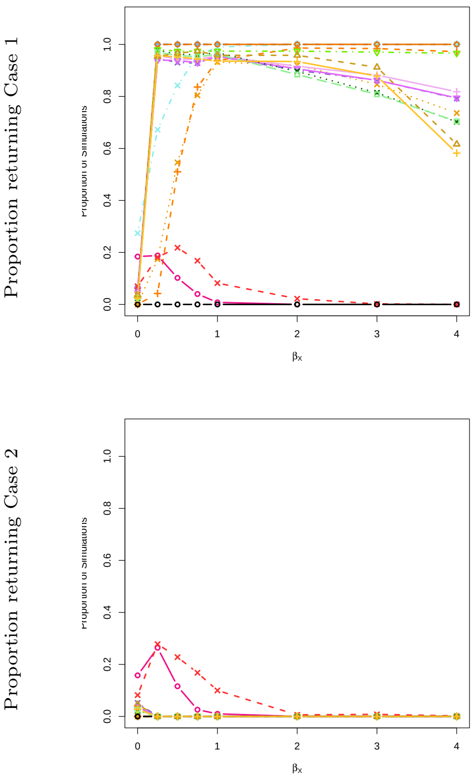

Proportion returning Case 1
Proportion returning Case 2
0.0
0.2
0.4
0.6
0.8
1.0
0
1
2
3
4
$\beta_x$

**(D.2)**

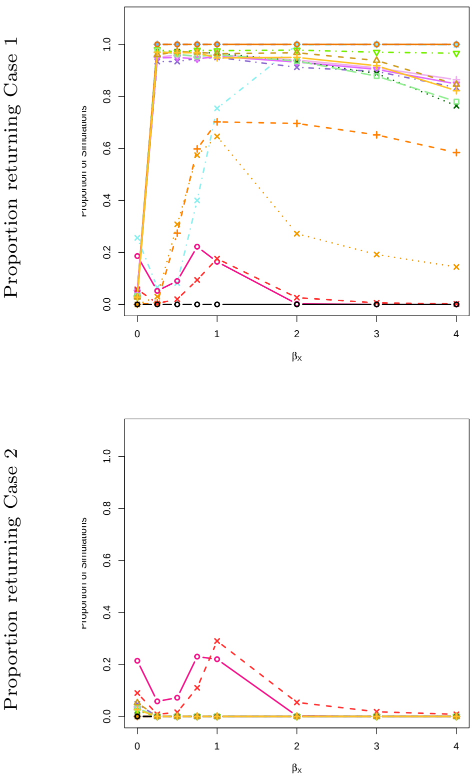

Proportion returning Case 1
Proportion returning Case 2
0.0
0.2
0.4
0.6
0.8
1.0
0
1
2
3
4
$\beta_x$

**(D.3)**

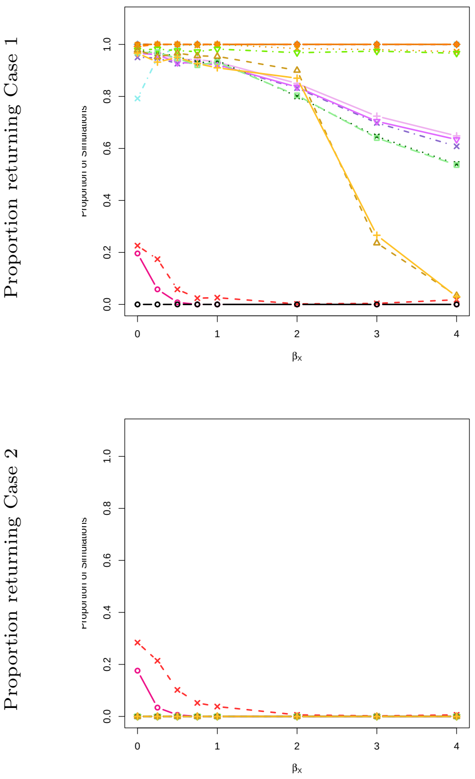

Proportion returning Case 1
Proportion returning Case 2
0.0
0.2
0.4
0.6
0.8
1.0
0
1
2
3
4
$\beta_x$

**(E)**

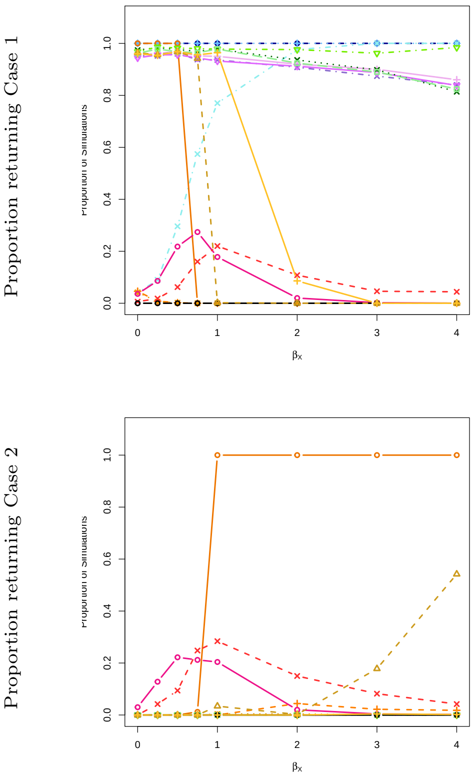

Proportion returning Case 1
Proportion returning Case 2
0.0
0.2
0.4
0.6
0.8
1.0
0
1
2
3
4
$\beta_x$

**(F)**

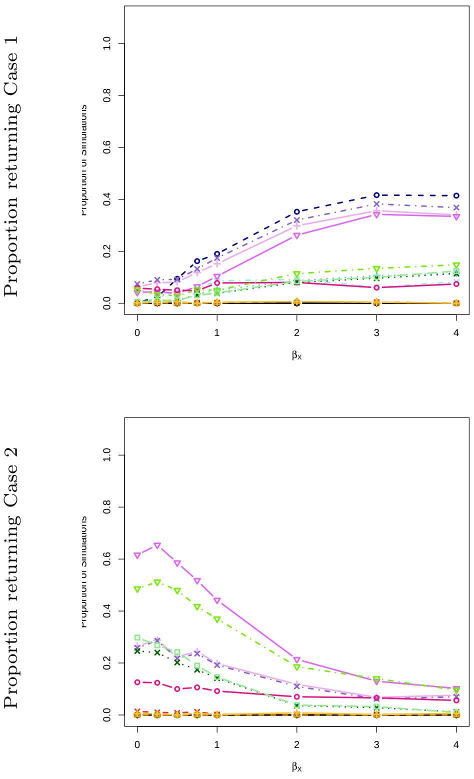

Proportion returning Case 1
Proportion returning Case 2
0.0
0.2
0.4
0.6
0.8
1.0
0
1
2
3
4
$\beta_x$

### 7 Phenotype 1: Bernoulli distributed; Phenotype 2: Bernoulli distributed; sample size $N = 100{,}000$; SNP effect size of 0.2.

**Supplemental Table 7.** Simulation results with $N = 100{,}000$ and 50 SNPs ($\gamma_{GX} = 0.2$, $\beta_{GY} = 0.2$). Heat maps summarize performance across the range of $\beta_X$ values shown in Supplemental Figure 7. Green cells indicate consistently high performance across all checks (no violations detected) in at least one setting. Otherwise, overlaid stripes show specific violations: inflated type I error ($> 10\%$, red), spurious $Y \to X$ inference ($> 5\%$, orange), or low power to detect $X \to Y$ ($< 75\%$, blue).

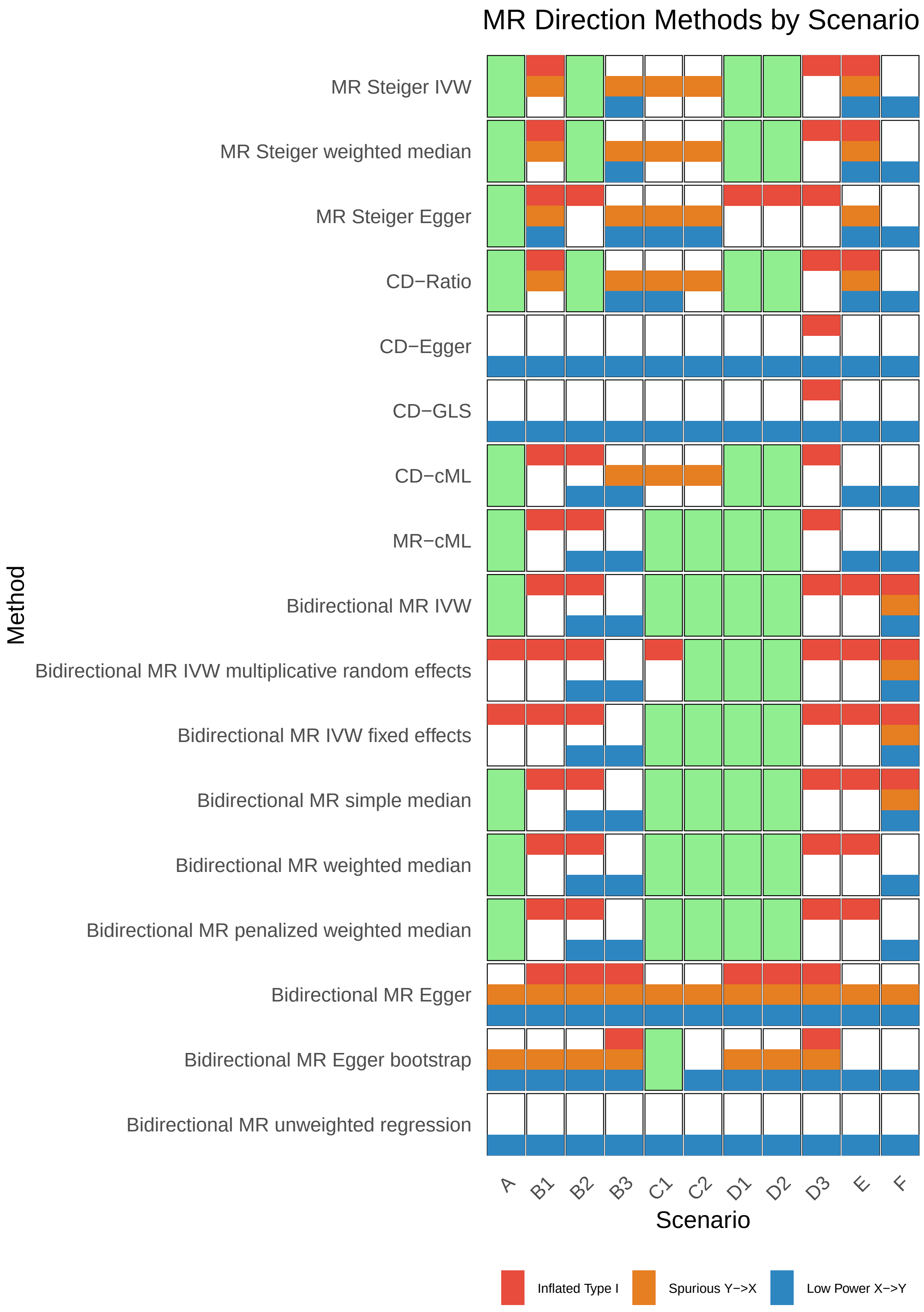

**Supplemental Figure 7.** Simulation results for Scenario 8 ($N$ = 100,000, 50 SNPs, $\gamma_{GX}$ = 0.2, $\beta_{GY}$ = 0.2). Both phenotypes are binary (Bernoulli distributed). Columns correspond to sub-scenarios (A–F); rows show the proportion of simulations returning case 1 ($X \to Y$, top) or case 2 ($Y \to X$, bottom) across values of $\beta_X$. The $x$-axis shows the true effect size $\beta_X$, and the $y$-axis shows the proportion of simulations where each method returned the given case. Line colors correspond to MR methods (see overview legend).

**(A)**

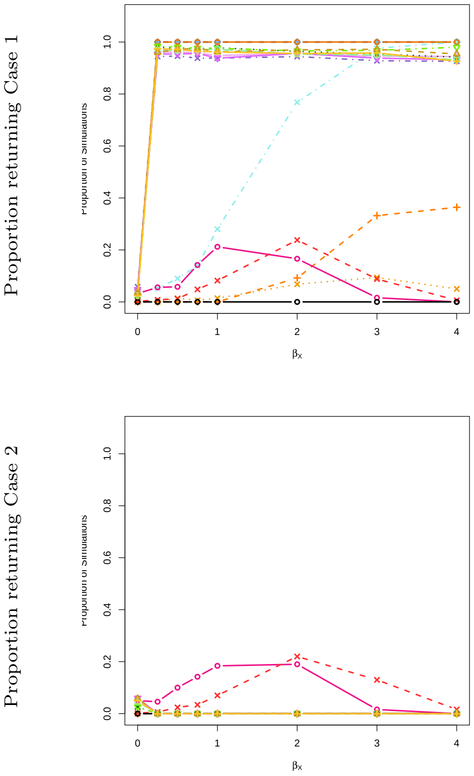

**(B.1)**

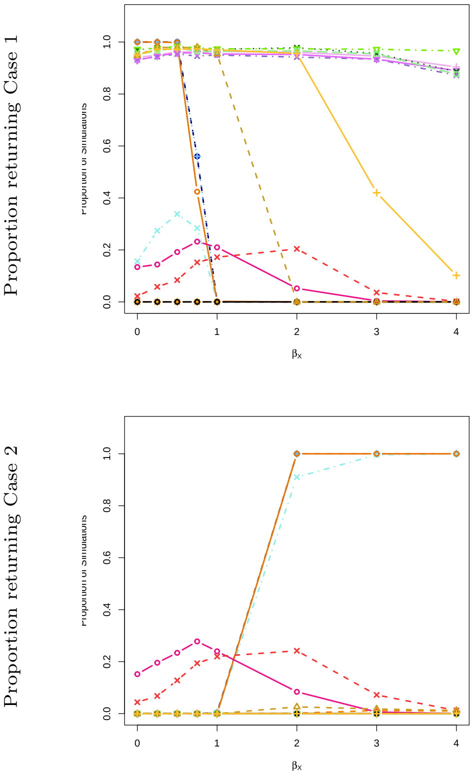

**(B.2)**

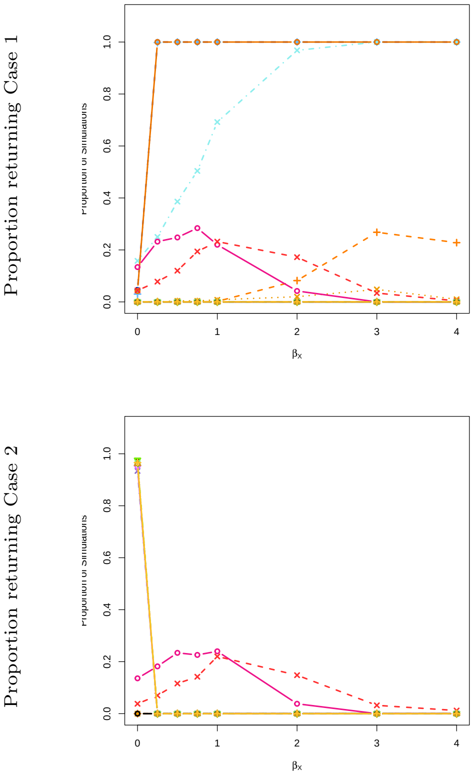

**(B.3)**

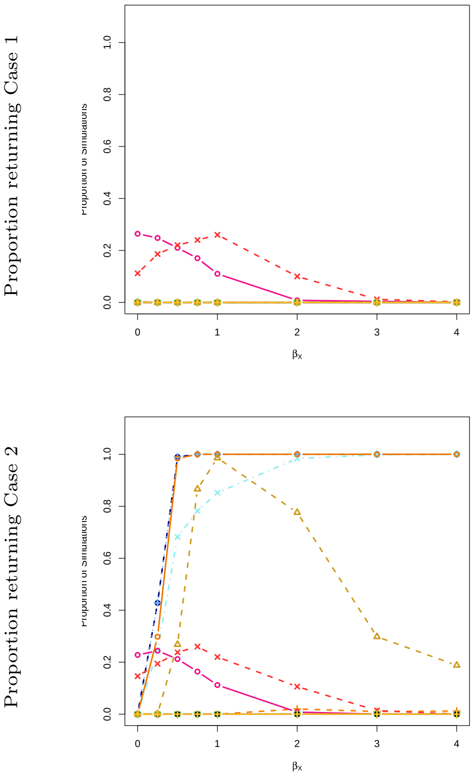

**(C.1)**

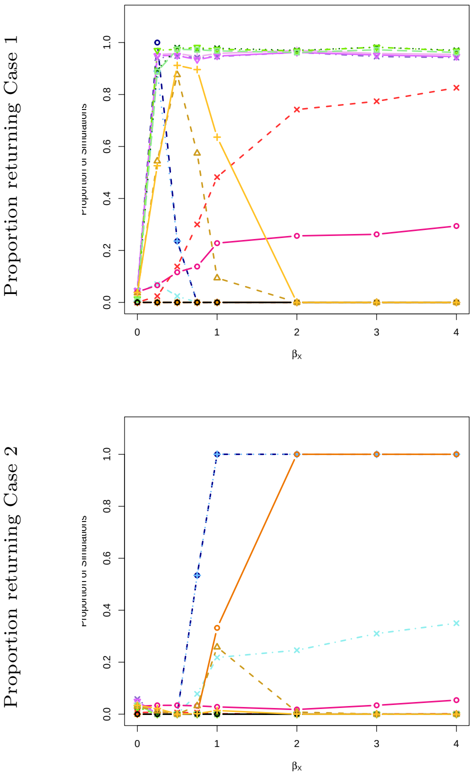

**(C.2)**

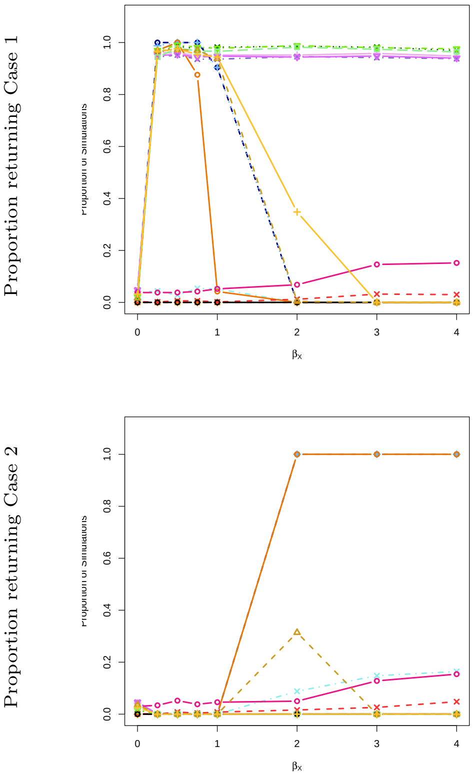

**(D.1)**

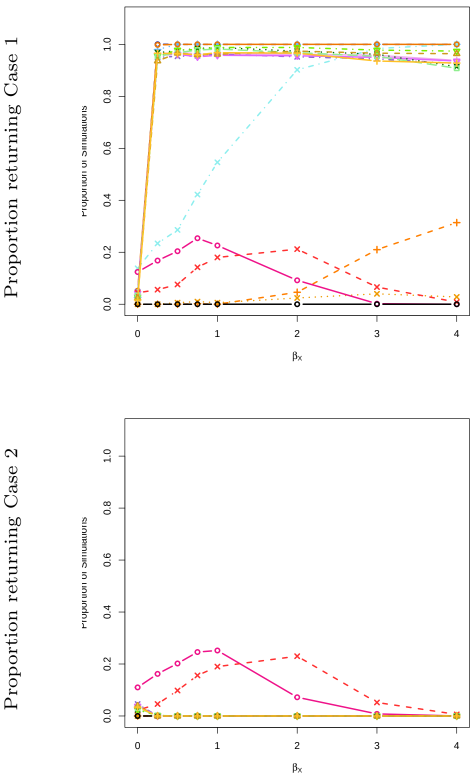
Proportion returning Case 1
Proportion returning Case 2
βx

**(D.2)**

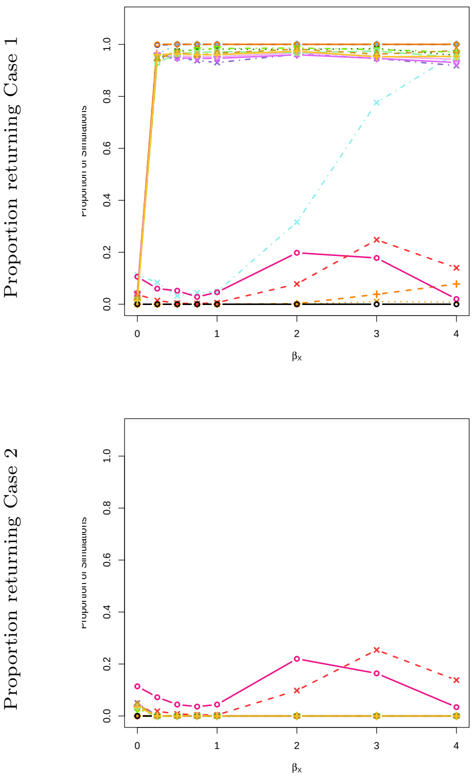
Proportion returning Case 1
Proportion returning Case 2
βx

**(D.3)**

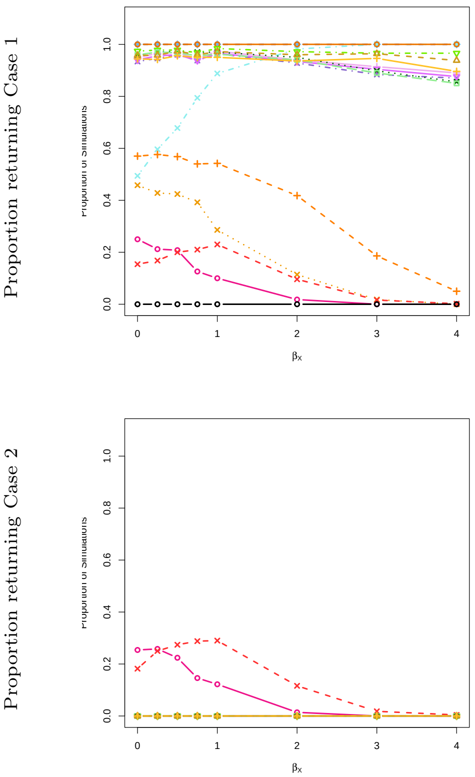
Proportion returning Case 1
Proportion returning Case 2
βx

**(E)**

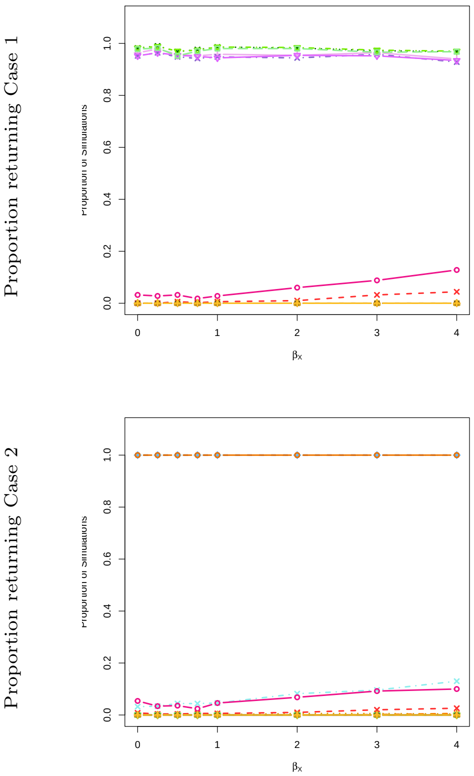
Proportion returning Case 1
Proportion returning Case 2
βx

**(F)**

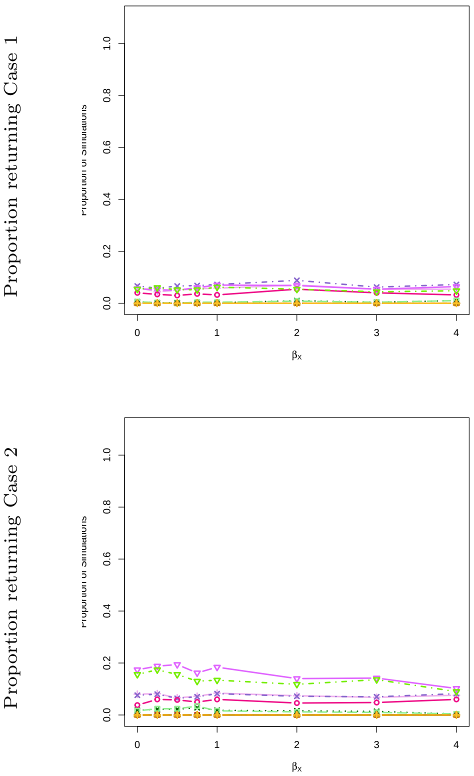
Proportion returning Case 1
Proportion returning Case 2
βx
